# Semaglutide is associated with stiffness improvement and broad liver benefits with distinct dose- and weight-linked patterns

**DOI:** 10.64898/2026.04.14.26350891

**Authors:** A.J. Venkatakrishnan, K Purushotham, Gowtham Varma, Avinash Aman, Karthik Murugadoss, Venky Soundararajan

## Abstract

Semaglutide has shown benefit in metabolic dysfunction-associated steatohepatitis (MASH), but real-world evidence across longitudinal liver phenotypes remains limited, particularly regarding how liver remodeling relates to weight loss and dose exposure. Using a de-identified federated electronic health record network spanning more than 29 million patients in the United States, including 489,785 semaglutide-treated adults, we analyzed 6,734 patients with baseline liver disease burden. We find that higher attained pre-landmark (0-2 years) semaglutide dose was associated with lower post-landmark (2-4 years) risk of steatohepatitis, alcoholic liver disease, and all-cause mortality, whereas greater pre-landmark weight loss was associated with lower post-landmark risk of steatohepatitis, steatotic liver disease, and hepatorenal syndrome, indicating distinct dose- and weight-linked patterns of long-term liver benefits. These associations were notable because semaglutide prescribing was generally lower during the post-landmark period, raising the possibility of durable benefit beyond peak exposure. Towards better understanding mechanistic bases for liver protection, we performed a complementary longitudinal study of 326 adults with paired noninvasive liver elastography measurements before and after treatment initiation. Median liver stiffness decreased from 4.85 [3.02 - 7.20] to 3.9 [2.6 - 5.8] kPa after semaglutide initiation (median change = −0.38 kPa; p<0.001), with 194 of 326 patients (59.5%) showing lower follow-up stiffness. A clinically meaningful reduction of at least 20% was observed in 133 of 326 patients (40.8%), and 69 of 326 (21.2%) shifted to a lower fibrosis stage by prespecified elastography thresholds. Larger improvements were also seen in patients with higher baseline stiffness (p<0.001); notably 80% of patients with cirrhosis-range baseline stiffness (≥12.5 kPa) achieved ≥20% improvement versus 29.5% with minimal baseline disease (p <0.001). The proportion achieving at least 20% stiffness improvement was similar across weight-loss strata, including patients with no weight loss or weight gain and those with at least 10% weight loss (38.0% in each group), and liver stiffness change showed negligible correlation with changes in weight, BMI, HBA1c, alanine aminotransferase, or aspartate aminotransferase. To provide biological context, single cell RNA analyses demonstrated sparse overall hepatic GLP1R expression (0.0239%), with enrichment in non-parenchymal niches including cholangiocytes, intrahepatic cholangiocytes, liver sinusoidal endothelial cells, and hepatic stellate cells implicated in fibrogenesis and vascular remodeling. Together, this real-world evidence suggests diverse liver benefits for semaglutide beyond weight-loss with intricate dose response relationships.

## Introduction

Metabolic dysfunction-associated steatotic liver disease (MASLD) is now the most common liver disease in adults and is closely linked to obesity, type 2 diabetes, and insulin resistance^1–4^. Across chronic liver diseases, fibrosis stage is the strongest predictor of liver-related complications and mortality, making fibrosis regression a central therapeutic goal^1–4^. Yet outside clinical trials, it remains difficult to document structural liver improvement longitudinally in routine practice.

The therapeutic landscape in MASLD and metabolic dysfunction-associated steatohepatitis (MASH) has changed rapidly. Resmetirom became the first US Food and Drug Administration-approved therapy for noncirrhotic MASH with moderate-to-advanced fibrosis in 2024^4,5^, and semaglutide subsequently showed histologic benefit in phase 3 MASH and received US regulatory approval for MASH with moderate-to-advanced fibrosis in 2025^6–9^. However, most available data still come from controlled trial populations with standardized follow-up, fixed dosing schedules, and protocolized endpoints. How semaglutide performs in ordinary care, where patients have heterogeneous comorbidity burdens, scan intervals, and treatment trajectories, is less well described.

Although MASLD and MASH provide the dominant clinical context for semaglutide-associated liver benefit, patients in routine care present with a broader spectrum of hepatic phenotypes and complications, including steatohepatitis, steatotic liver disease, cirrhosis, portal hypertensive complications, alcohol-associated liver disease, hepatorenal syndrome, cholestatic presentations, viral hepatitis, hepatocellular carcinoma, and liver-related mortality. Whether semaglutide-associated benefit is uniform across this heterogeneous landscape, or instead shows distinct relationships with treatment intensity and weight loss across different liver phenotypes, remains poorly understood.

Liver elastography is widely used in clinical practice as a noninvasive estimate of liver stiffness and, by extension, fibrosis burden. Although stiffness is not identical to histologic fibrosis and can be influenced by inflammation, congestion, or technical factors, serial liver stiffness measurement (LSM) has prognostic value and captures clinically relevant disease progression or regression over time^10,11^. In real-world hepatology, many of these measurements are documented in semi-structured or free-text radiology reports rather than in analysis-ready tables, creating a practical data-extraction bottleneck. At the same time, the cellular targets through which GLP-1 receptor agonism may influence hepatic remodeling remain incompletely understood.

Beyond structural fibrosis measures, liver disease progression and clinical outcomes in MASLD are influenced by multiple, partially overlapping pathways, including steatohepatitis activity, metabolic remodeling, and systemic inflammatory signaling. Emerging data suggest that therapeutic effects of GLP-1 receptor agonists may not be uniform across these domains, with some outcomes potentially more closely linked to weight loss, while others may reflect dose-dependent or weight-independent mechanisms. However, these relationships remain poorly characterized in real-world populations, where longitudinal imaging, treatment intensity, and downstream clinical outcomes can be evaluated together.

Here, we used an agentic large language model (LLM) workflow to reconstruct longitudinal elastography, treatment exposure, and liver outcomes from structured and unstructured electronic health record (EHR) data within a large-scale de-identified federated network. We sought to (1) characterize longitudinal changes in elastography-derived liver stiffness after semaglutide initiation; (2) evaluate dose- and weight-linked patterns across downstream liver outcomes in routine care; and (3) assess whether observed structural and clinical liver benefits follow shared or distinct pathways in real-world populations.

## Results

### Dose-escalation was associated with greater weight loss as well as lower post-landmark risk of steatohepatitis, alcoholic liver disease, and all-cause mortality

Of 269,390 patients who initiated semaglutide between March 2018 and January 2024, 6,734 had baseline liver conditions and constituted the cohort examined in this observational study (**Table 1** and **Table S1**). Among 6,734 semaglutide-treated patients with baseline liver burden, 6,051 met eligibility for the 2-year post-landmark dose analysis: requiring an analyzable maximum dose within low (0.25–1.0 mg, n=4,218) or high (≥1.7 mg, n=1,833) dose bins and continued follow-up beyond the 2-year landmark. In this subset, maximum body-weight reduction deepened progressively with increasing semaglutide dose across the full 0.25–2.4 mg range, following an approximately linear gradient corresponding to an estimated 3.08% greater maximum weight loss per 1 mg increase in attained dose (**Fig. 1**). Three hepatic outcomes showed nominally significant differences by maximum dose group over the 2-year landmark period (**Fig. 2**). Incident steatohepatitis (MASH/NASH) was the strongest dose-dependent signal, with a lower 2-year event rate in the high-dose group compared with low dose (3.5% vs. 5.5%; RR=0.64, p=0.026). Incident alcoholic liver disease similarly favoured high-dose semaglutide (0.8% vs. 1.7%; RR=0.48, p=0.025), as did all-cause mortality (4.0% vs. 5.8%; RR=0.68, p=0.039). All three effects were directionally consistent with dose-dependent hepatoprotection, though log-rank curve separation was modest for alcoholic liver disease and mortality, suggesting the absolute event-rate contrast was more apparent than the full temporal divergence across follow-up. The remaining 17 endpoints, including cirrhosis, ascites, portal hypertension, hepatic encephalopathy, liver fibrosis, hepatocellular carcinoma, and MASLD/NAFLD did not significantly differ by dose group (all p>0.10; **Table 2**).

**Fig. 1.**
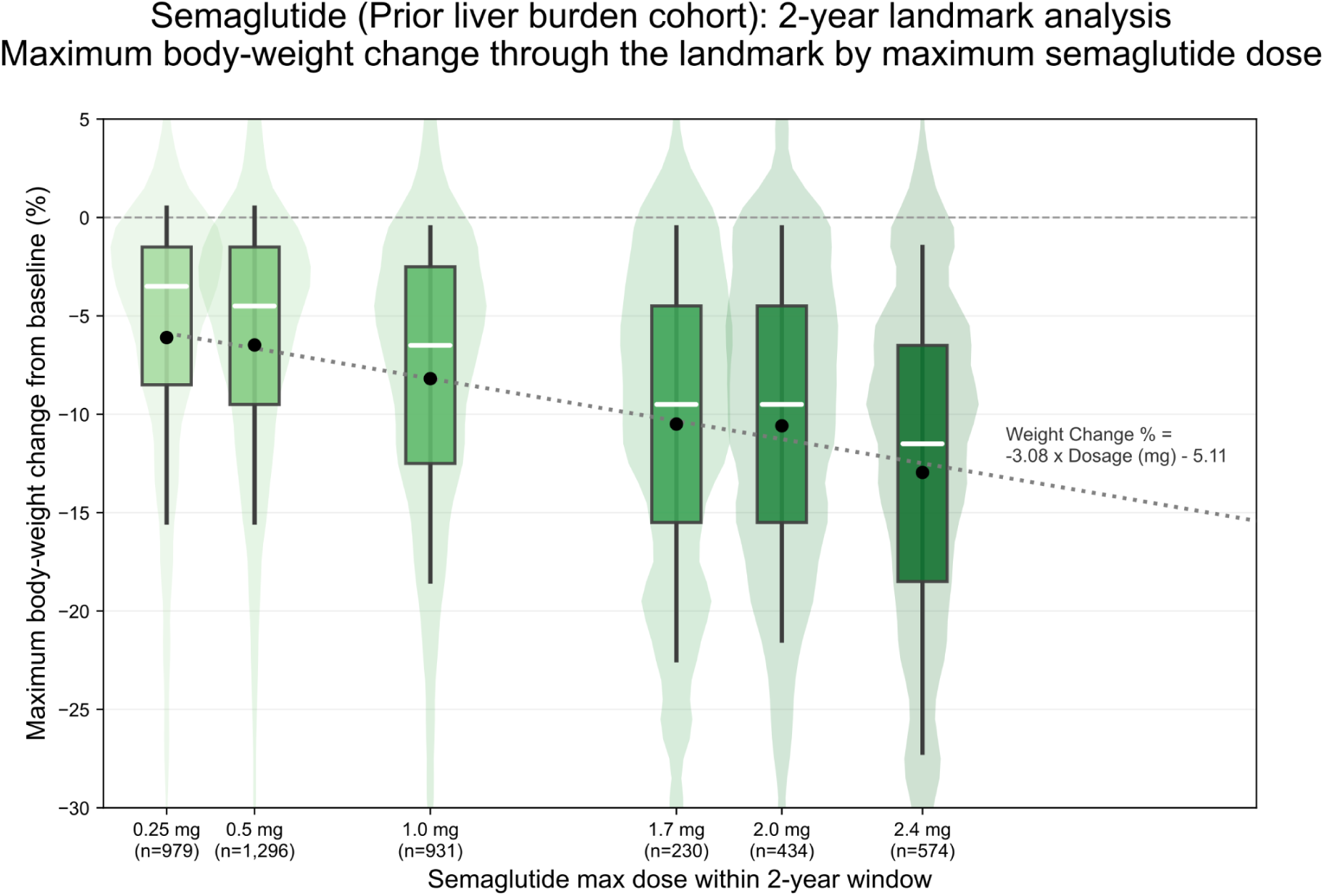
Relationship between semaglutide maximum dose and maximum weight loss through the 2-year landmark date in the prior liver burden cohort. Boxplots show the distribution of maximum percent body-weight change from baseline achieved within the 2-year landmark window, stratified by the maximum semaglutide dose attained during that period. Weight loss is represented as negative values. Center lines denote medians, boxes span the interquartile range, whiskers reflect the overall data spread, and black dots mark group means. The dotted line represents the fitted linear relationship between dose and weight change, with the regression equation displayed on the plot. The number of patients in each dose category is shown below the x-axis.

**Fig. 2.**
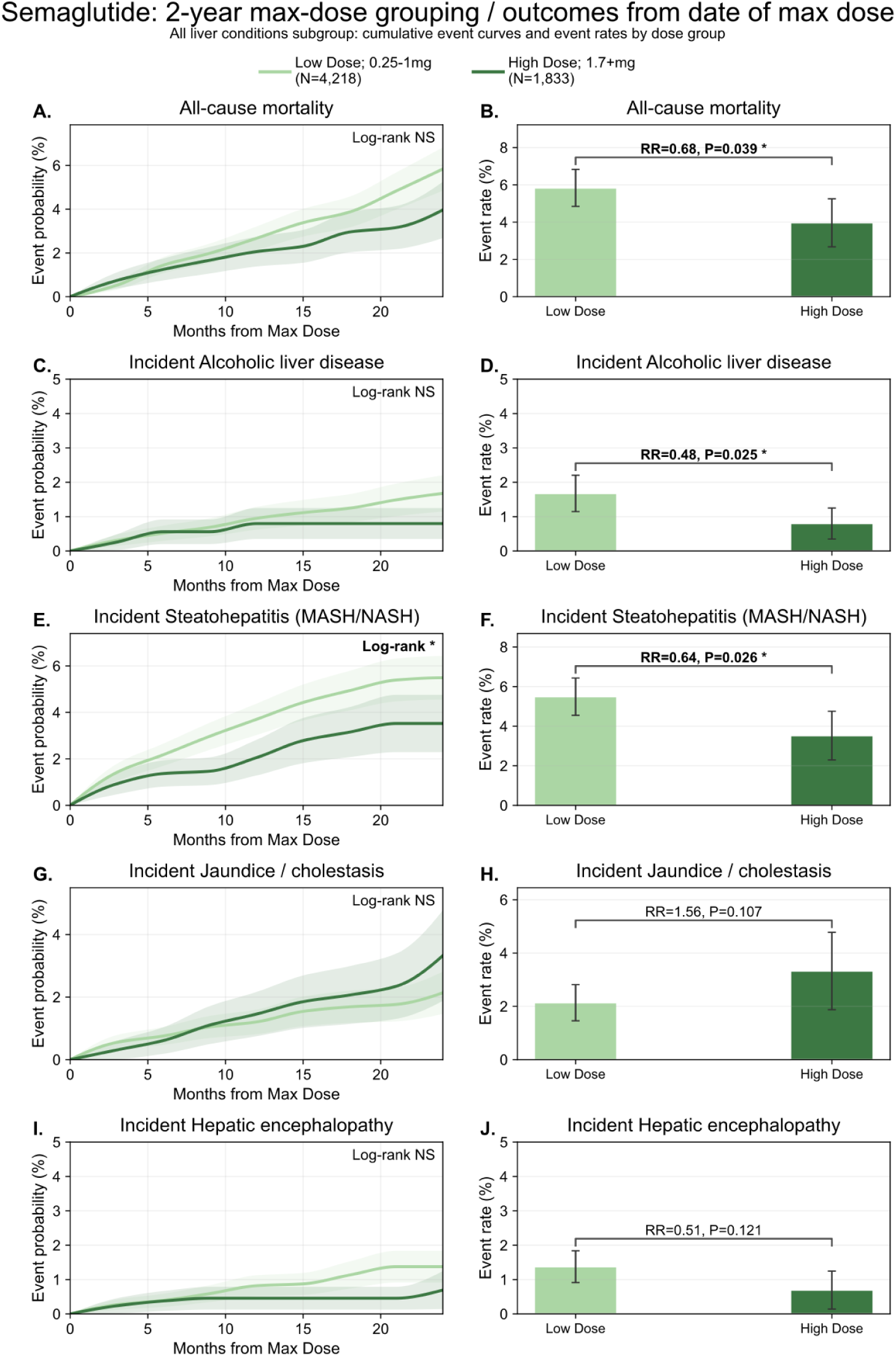
Higher maximum semaglutide dose was associated with lower post-landmark liver disease risk in patients with prior liver burden. Patients were grouped by the maximum semaglutide dose reached by the 2-year landmark as low dose (0.25-1.0 mg; n = 4,218) or high dose (≥1.7 mg; n = 1,833). Left-column panels show cumulative event curves after the landmark and right-column panels show corresponding post-landmark event rates for **(A-B)** all-cause mortality, **(C-D)** incident alcoholic liver disease, **(E-F)** incident steatohepatitis (MASH/NASH), **(G-H)** incident jaundice/cholestasis, and **(I-J)** incident hepatic encephalopathy. Relative risks and nominal P values are shown on the bar plots. Higher-dose semaglutide was associated with significantly lower risk of all-cause mortality, incident steatohepatitis, and incident alcoholic liver disease.

**Table 1.**
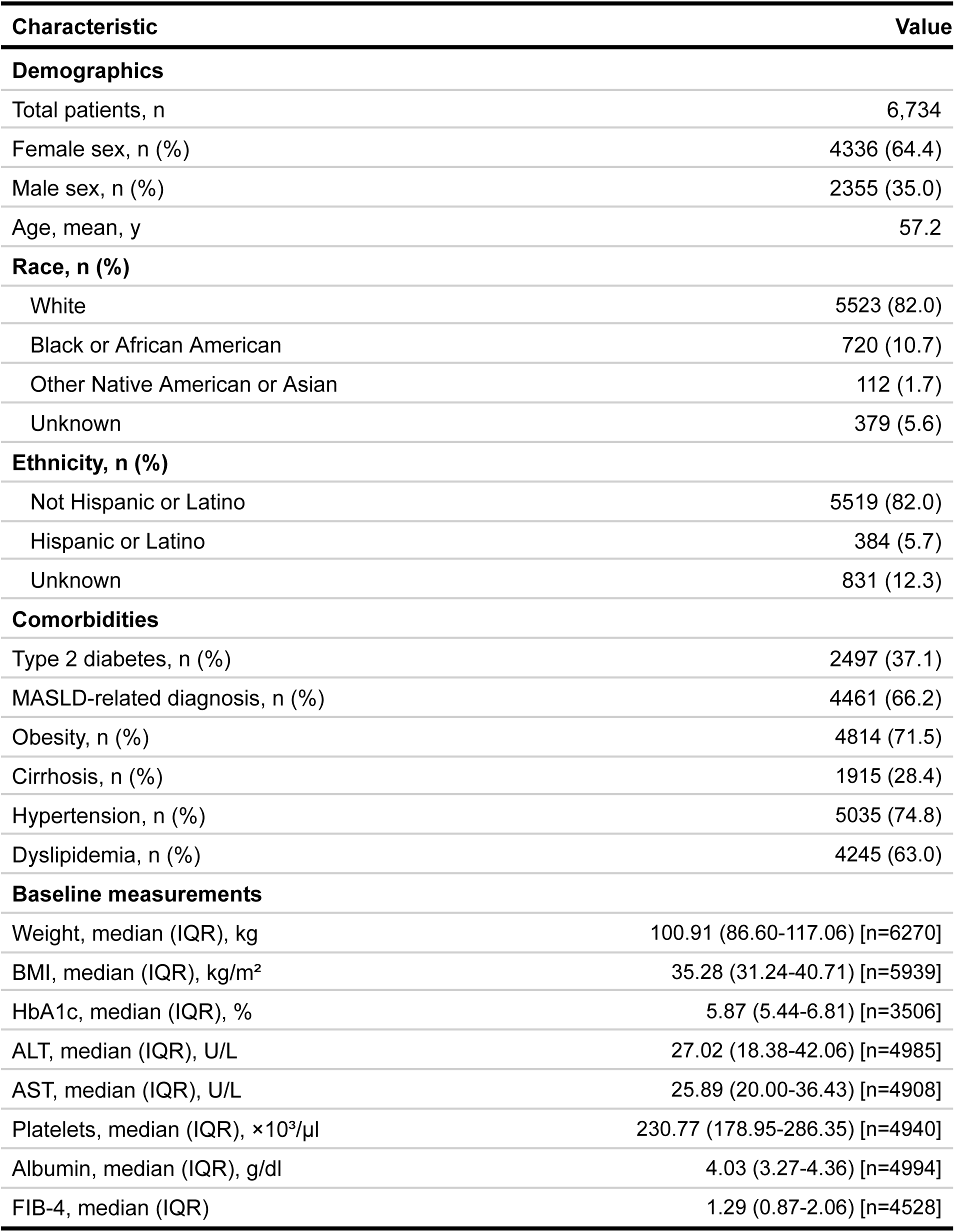
Cohort demographics and baseline characteristics.

| Characteristic | Value |
| --- | --- |
| **Demographics** |  |
| Total patients, n | 6,734 |
| Female sex, n (%) | 4336 (64.4) |
| Male sex, n (%) | 2355 (35.0) |
| Age, mean, y | 57.2 |
| **Race, n (%)** |  |
| White | 5523 (82.0) |
| Black or African American | 720 (10.7) |
| Other Native American or Asian | 112 (1.7) |
| Unknown | 379 (5.6) |
| **Ethnicity, n (%)** |  |
| Not Hispanic or Latino | 5519 (82.0) |
| Hispanic or Latino | 384 (5.7) |
| Unknown | 831 (12.3) |
| **Comorbidities** |  |
| Type 2 diabetes, n (%) | 2497 (37.1) |
| MASLD-related diagnosis, n (%) | 4461 (66.2) |
| Obesity, n (%) | 4814 (71.5) |
| Cirrhosis, n (%) | 1915 (28.4) |
| Hypertension, n (%) | 5035 (74.8) |
| Dyslipidemia, n (%) | 4245 (63.0) |
| **Baseline measurements** |  |
| Weight, median (IQR), kg | 100.91 (86.60-117.06) [n=6270] |
| BMI, median (IQR), kg/m<sup>2</sup> | 35.28 (31.24-40.71) [n=5939] |
| HbA1c, median (IQR), % | 5.87 (5.44-6.81) [n=3506] |
| ALT, median (IQR), U/L | 27.02 (18.38-42.06) [n=4985] |
| AST, median (IQR), U/L | 25.89 (20.00-36.43) [n=4908] |
| Platelets, median (IQR), ×10<sup>3</sup>/μl | 230.77 (178.95-286.35) [n=4940] |
| Albumin, median (IQR), g/dl | 4.03 (3.27-4.36) [n=4994] |
| FIB-4, median (IQR) | 1.29 (0.87-2.06) [n=4528] |

**Table 2.** Liver-related outcomes: event counts, relative risk by maximum achieved dose, and associations with maximum achieved dose and weight loss. Green shading indicates nominally significant associations with maximum achieved dose (p < 0.05), and yellow shading indicates nominally significant associations with maximum achieved weight loss (p < 0.05). Outcomes marked with * were significantly associated with maximum achieved dose, whereas outcomes marked with ⇑ were significantly associated with maximum achieved weight loss. Values <11 were suppressed because of low event counts; N/A indicates not applicable.

| Outcome | Number of patients at risk | Number of patients with event/outcome | Rate ratio: High dose vs Low | P-value (High Dose vs Low Dose) | P-value (High vs Low Weight Loss) |
| --- | --- | --- | --- | --- | --- |
| **All-cause mortality\*** | 6051 | 166 | 0.68 | **0.0388** | 0.1348 |
| **Incident Alcoholic liver disease\*** | 5674 | 47 | 0.48 | **0.0248** | 0.4638 |
| **Alcoholic Steatohepatitis\*↑ (MASH/NASH)** | 5050 | 155 | 0.64 | **0.0255** | **0.0055** |
| Jaundice / cholestasis | 4671 | 62 | 1.56 | 0.1074 | 0.4056 |
| Hepatic encephalopathy | 5858 | 40 | 0.51 | 0.1208 | 0.062 |
| Liver fibrosis | 5787 | 109 | 0.74 | 0.1425 | 0.6592 |
| **Viral hepatitis C↑** | 5676 | $<11$ | 2.42 | 0.1586 | **0.017** |
| Cirrhosis | 4882 | 143 | 1.26 | 0.1811 | 0.2806 |
| **Incident Steatotic liver disease (MASLD/NAFLD)↑** | 2941 | 244 | 0.83 | 0.1844 | **0.0158** |
| Hepatocellular carcinoma | 5894 | 39 | 1.47 | 0.2333 | 0.2658 |
| Drug-induced liver injury | 6029 | $<11$ | 2.42 | 0.273 | 0.5563 |
| Spontaneous bacterial peritonitis | 6025 | $<11$ | 1.81 | 0.3842 | 0.1685 |
| **Hepatorenal syndrome↑** | 6009 | 19 | 0.61 | 0.386 | **0.0004** |
| Autoimmune / cholestatic liver disease | 5751 | 25 | 0.79 | 0.5909 | 0.3727 |
| Portal hypertension | 5566 | 159 | 1.08 | 0.6414 | 0.295 |
| Acute liver failure | 5756 | 66 | 0.88 | 0.651 | 0.4606 |
| Ascites | 5624 | 112 | 1.04 | 0.863 | 0.534 |
| Esophageal varices | 5643 | 114 | 0.97 | 0.8963 | 0.1909 |
| Liver transplant | 6048 | $<11$ | 0 | N/A | N/A |
| Viral hepatitis B | 5953 | $<11$ | 0 | N/A | 0.6744 |

### Greater weight loss during the landmark period was associated with lower post-landmark risk of steatohepatitis and steatotic liver disease

Patients were categorised by maximum weight loss achieved before the landmark as <5% (n=2,129), 5–10% (n=1,150), 10–15% (n=715), 15–20% (n=411), or ≥20% (n=442). Incident steatohepatitis differed significantly across weight-loss strata (global log-rank p=0.006, **Fig. 3**), though the relationship was not monotonic. The highest event rate appeared in the 5–10% weight-loss group, with lower rates at greater weight loss. Incident steatotic liver disease (MASLD/NAFLD) also varied significantly across strata (global log-rank p=0.016), with a broadly protective gradient at higher weight loss. Incident hepatorenal syndrome showed the strongest statistical weight-loss signal (global log-rank p=0.0004, **Table 2**), though the small event count warrants caution in interpretation. All remaining outcomes, including liver fibrosis, cirrhosis, ascites, portal hypertension, hepatic encephalopathy, acute liver failure, esophageal varices, hepatocellular carcinoma, jaundice/cholestasis, autoimmune/cholestatic liver disease, spontaneous bacterial peritonitis, drug-induced liver injury, viral hepatitis B and C, and liver transplant, showed no significant differences across weight-loss strata (all p>0.10; **Table 3**), with several rare outcomes having fewer than 10 total events.

**Fig. 3.**
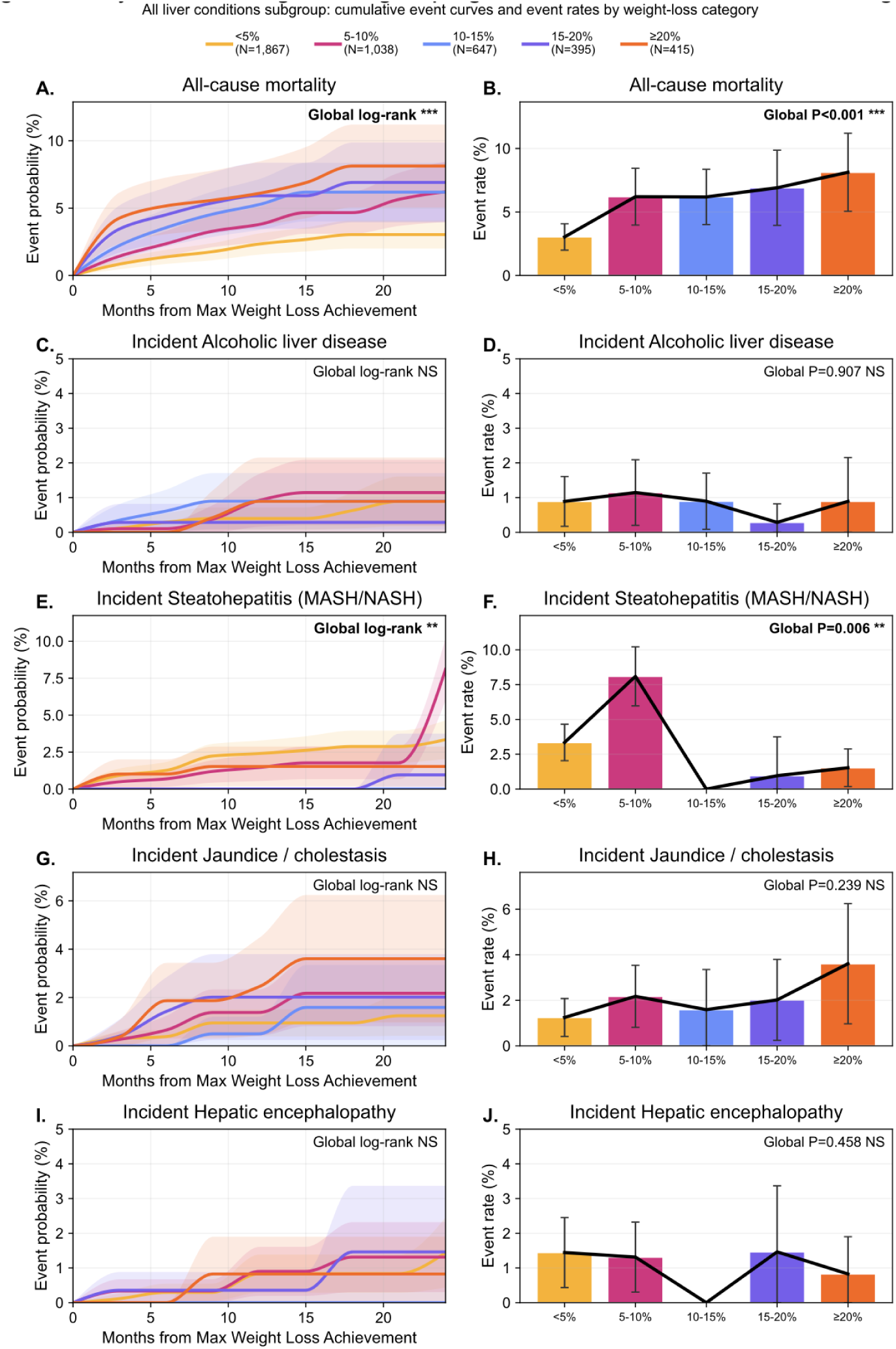
Greater weight loss was associated with lower post-landmark risk of steatohepatitis in the prior liver burden cohort. Patients were categorized according to the maximum percentage reduction in body weight achieved before the 2-year landmark as <5% (n = 2,129), 5–10% (n = 1,150), 10–15% (n = 715), 15–20% (n = 411), or ≥20% (n = 442), and liver outcomes were assessed only after the landmark among patients event-free through that timepoint. Left-column panels show cumulative post-landmark event probability over follow-up for all-cause mortality (A), incident alcoholic liver disease (C), incident steatohepatitis/MASH (E), incident jaundice/cholestasis (G), and incident hepatic encephalopathy (I). Right-column panels show the corresponding post-landmark event rates across weight-loss categories for all-cause mortality (B), incident alcoholic liver disease (D), incident steatohepatitis/MASH (F), incident jaundice/cholestasis (H), and incident hepatic encephalopathy (J), with global P values indicated. Greater weight loss was significantly associated with lower post-landmark risk of steatohepatitis (global P = 0.006), with a trend observed for hepatic encephalopathy (P = 0.062); the remaining outcomes in this panel did not show significant weight-loss gradients.

**Table 3.** Cohort demographics and baseline characteristics for patients with serial elastography.

| Characteristic | Value |
| --- | --- |
| **Demographics** |  |
| Total patients, n | 326 |
| Female sex, n (%) | 185 (56.7) |
| Male sex, n (%) | 141 (43.3) |
| Age, mean, y | 53.8 |
| **Race, n (%)** |  |
| White | 284 (87.1) |
| Black or African American | 16 (4.9) |
| Other Native American or Asian | 17 (2.8) |
| Unknown | 9 (2.8) |
| **Ethnicity, n (%)** |  |
| Not Hispanic or Latino | 274 (84.0) |
| Hispanic or Latino | 45 (13.8) |
| Unknown | 7 (2.1) |
| **Comorbidities** |  |
| Type 2 diabetes, n (%) | 209 (64.1) |
| MASLD-related diagnosis, n (%) | 225 (69.0) |
| ICD coded | 158 (48.5) |
| NLP extracted | 192 (58.9) |
| Obesity, n (%) | 254 (77.9) |
| Cirrhosis, n (%) | 196 (60.1) |
| Hypertension, n (%) | 253 (77.6) |
| Dyslipidemia, n (%) | 237 (72.7) |
| **Baseline measurements** |  |
| Liver stiffness, median (IQR), kPa | 4.85 (3.02-7.20) [n=326] |
| Weight, median (IQR), kg | 93.10 (81.12-108.03) [n=295] |
| BMI, median (IQR), kg/m<sup>2</sup> | 33.70 (30.70-36.10) [n=257] |
| HbA1c, median (IQR), % | 6.40 (5.70-7.80) [n=236] |
| ALT, median (IQR), U/L | 36.00 (22.00-60.50) [n=303] |
| AST, median (IQR), U/L | 31.00 (21.00-47.00) [n=301] |
| Platelets, median (IQR), ×10<sup>3</sup>/μl | 219.00 (162.00-268.00) [n=298] |
| Albumin, median (IQR), g/dl | 4.40 (4.20-4.60) [n=288] |
| FIB-4, median (IQR) | 1.31 (0.94-2.69) [n=290] |
| **Baseline fibrosis severity, n (%)** |  |
| Minimal (<7 kPa) | 234 (71.8) |
| Significant (7-9.5 kPa) | 39 (12.0) |
| Advanced (9.5-12.5 kPa) | 23 (7.1) |
| Cirrhosis-range ($\geq 12.5$ kPa) | 30 (9.2) |

Data are presented as n (%), median (IQR), or mean, as indicated. Percentages were calculated using the full cohort as the denominator. For baseline laboratory and anthropometric measures, the contributing sample size is shown in brackets. MASLD-related diagnosis reflects NAFLD/NASH identified by ICD coding or natural language processing. MASLD denotes metabolic dysfunction-associated steatotic liver disease; NLP, natural language processing; IQR, interquartile range; FIB-4, fibrosis-4 index.

**Table 4.** Paired elastography outcomes in the primary analytic cohort.

| Outcome | Value |
| --- | --- |
| Paired analytic cohort, n | 326 |
| Baseline liver stiffness, median (IQR), kPa | 4.85 (3.02-7.20) |
| Follow-up liver stiffness, median (IQR), kPa | 3.90 (2.60-5.80) |
| Within-patient change in liver stiffness, median (IQR), kPa | -0.38 (-2.23 to 0.67) |
| Wilcoxon signed-rank P value | $8.98 \times 10^{-7}$ |
| Any decrease in stiffness, n (%) | 194 (59.5) |
| Any increase in stiffness, n (%) | 121 (37.1) |
| No change, n (%) | 11 (3.4) |
| At least 20% reduction in stiffness, n (%) | 133 (40.8) |
| Stage regression, n (%) | 69 (21.2) |
| Stage progression, n (%) | 30 (9.2) |
| Baseline scan timing, median days before index (IQR) | 209 (56-405) |
| Follow-up scan timing, median days after index (IQR) | 445 (315-711) |

Data are presented as median (IQR) or n (%), unless otherwise indicated. The primary paired comparison used the Wilcoxon signed-rank test because liver stiffness values were right-skewed. In descriptive sensitivity analyses, mean liver stiffness decreased from 6.49 (s.d. 6.56) kPa at baseline to 5.33 (s.d. 5.08) kPa at follow-up, with a mean within-patient change of -1.16 kPa; the paired t-test P value was $4.56 \times 10^{-3}$ . Baseline was defined as the closest pre-index stiffness measurement and follow-up as the latest post-index stiffness measurement within the prespecified analytic windows.

### Semaglutide therapy was associated with reduced liver stiffness

We analyzed 326 semaglutide-treated patients with available quantitative liver stiffness measurements (**Tables S2-8**) during the pre-index (at most 5 years before starting semaglutide) and post-index period (at least 6 months and at most 5 years after starting semaglutide). The baseline scan occurred on average 313 ± 338 days before semaglutide initiation, and the follow-up scan occurred on average 541±303 days after initiation. The mean age of this patient cohort was 53.8±6 years, and 185 of 326 patients (56.7%) were women. Regarding pre-existing comorbidities, 64.1% had type 2 diabetes, 77.9% were obese, and 69.0% had a MASLD-related diagnosis captured through legacy NAFLD/NASH coding or NLP extraction (**Table 3**). Median baseline liver stiffness was 4.85 kPa [IQR 3.02–7.20]. By prespecified thresholds (see **Methods**), 234 of 326 patients (71.8%) were in the F0–F1 range, 39 of 326 (12.0%) in F2, 23 of 326 (7.1%) in F3, and 30 or a 326 (9.2%) in the cirrhosis-range F4 category.

Across the full cohort with available high-confidence serial elastography data, median liver stiffness decreased from 4.85 [3.02 - 7.20] kPa before treatment to 3.9 [2.6 - 5.8] kPa after treatment, corresponding to a median within-patient change of 0.38 kPa (Wilcoxon signed-rank test = 8.98 × 10^−7^) (**Fig. 4a**). Directionally, 194 of 326 patients (59.5%) had lower follow-up stiffness, 121 of 326 (37.1%) had higher follow-up stiffness, and 11 of 326 (3.4%) were unchanged (**Fig. 4b**). A reduction of at least 20% was observed in 133 of 326 patients (40.8%). Using established elastography-based fibrosis thresholds, 69 of 326 patients (21.2%) showed stage regression whereas 30 of 326 (9.2%) showed stage progression (**Fig. 4c**). Among patients with baseline LSM values ≥7 kPa (n=92), consistent with significant fibrosis, 63% transitioned to <7kPa on follow-up, indicating regression. Among those with baseline values ≥9.5 kPa (n=53), consistent with advanced fibrosis, 72% decreased below this threshold after treatment. Among patients with a baseline LSM ≥12.5 kPa consistent with cirrhotic range disease (n=30), 77% showed a reduction in stiffness with LSM <12.5 kPa during the follow-up period Conversely, progression to a higher stage was observed in only 11% of patients with significant baseline fibrosis (≥7 kPa).

**Fig. 4.**
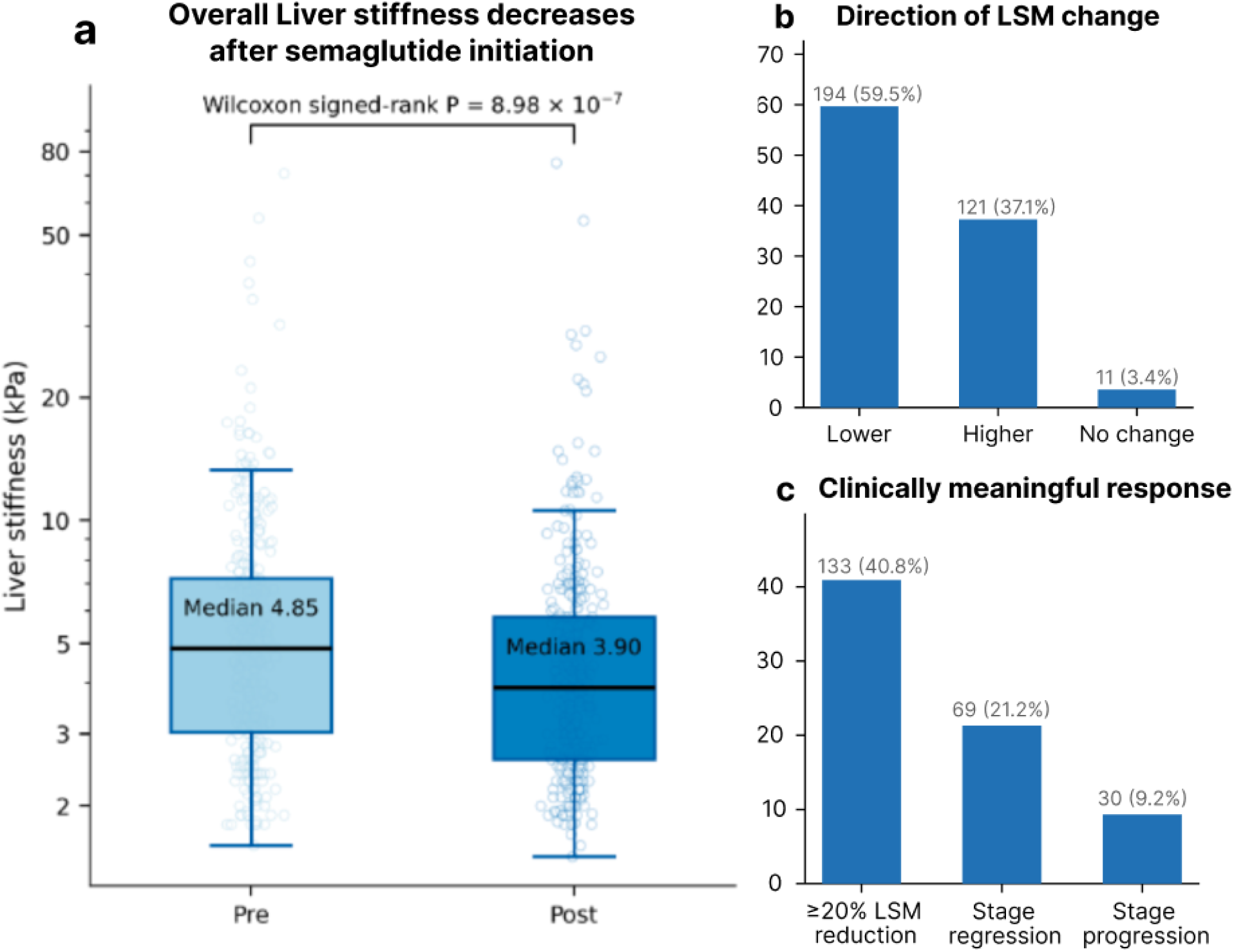
Cohort assembly and overall liver stiffness signal. **a.** Pre-index versus post-index liver stiffness with paired observations and summary distribution. **b.** Direction of within-patient change (decrease, increase, unchanged). **c.** Distribution of >=20% stiffness reduction and elastography-based stage regression/progression.

### The degree of liver stiffness reduction was largest in patients with the most severe baseline disease

We assessed whether there was a relationship between baseline liver stiffness and reduction in liver stiffness (**Fig. 5a-b**). Baseline liver stiffness was significantly associated with the change in liver stiffness following semaglutide initiation (P = 2.54 × 10^−19^). Patients with liver stiffness below 7.0 kPa at baseline showed minimal change after semaglutide initiation (mean change +0.42 kPa), whereas patients with baseline liver stiffness measurements of 7.0–9.5 kPa, 9.5–12.5 kPa, and ≥12.5 kPa had progressively larger median changes of −3 [−4.7, −0.35], −5.1 [−6.95, −2.83], and −9.4 [−14.7, −3.7] kPa, respectively (Kruskal–Wallis P = 4.21 × 10^−14^). Rates of clinically meaningful improvement (≥20% reduction in stiffness^12^) rose with worsening baseline severity: 29.5% in the <7 kPa group, 56.4% in the 7–9.5 kPa group, 78.3% in the 9.5–12.5 kPa group, and 80.0% in the ≥12.5 kPa group. Stage regression was evaluated among patients with baseline liver stiffness ≥7 kPa and occurred in 69.2%, 82.6%, and 76.7% among patients starting in the 7-9.5 kPa, 9.5-12.5 kPa, and ≥12.5 kPa groups, respectively. At the stage-transition level, 58 of 92 patients (63.0%) who began in F2–F4 were in the F0–F1 range at follow-up (**Fig. 5c**).

**Fig. 5.**
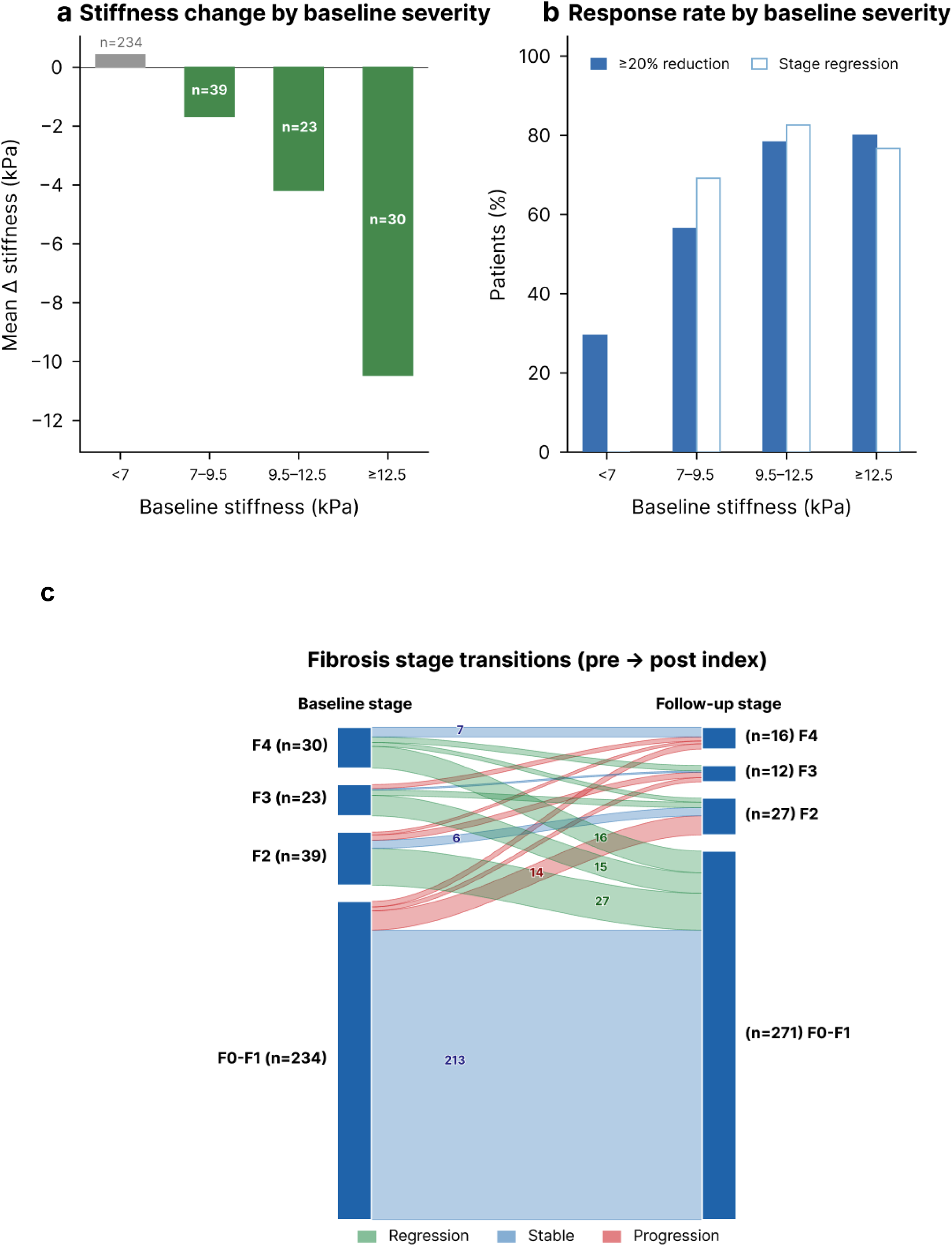
Greater improvement with worse baseline stiffness. Patients with higher baseline liver stiffness showed larger absolute reductions and higher rates of clinically meaningful improvement and stage regression. **a.** The left panel summarizes mean change in liver stiffness by baseline severity stratum. **b.** The right panel summarizes rates of >=20% reduction and stage regression. **c.** Sankey-style diagram showing movement between pragmatic elastography-based fibrosis categories from baseline to follow-up. Stages were defined as F0-F1 (<7.0 kPa), F2 (7.0-9.5 kPa), F3 (9.5-12.5 kPa), and F4/cirrhosis-range (>=12.5 kPa).

### Degree of liver stiffness reduction was not correlated with weight loss magnitude, changes in metabolic parameters, or baseline cardiometabolic comorbidities

We assessed whether there was a relationship between weight change and reduction in liver stiffness (**Fig. 6**). In the subset with repeated pre- and post-initiation body weight measurements (n = 250), weight decreased by a median of 4.85 [2.03, 12.55] kg and BMI decreased by a median of 1.80 [0.17, 3.78] kg/m². Interestingly, the fraction of patients achieving at least 20% stiffness reduction was similar across weight-loss strata: 39.7% with ≥10% weight loss, 50.0% with 5–10% weight loss, 38.3% with <5% weight loss, and 39.7% in patients with no weight loss or weight gain (**Fig. 6** and **Table S3**). Stage regression rates were also similar across weight change strata – 16.8% with ≥10% weight loss, 28.6% with 5–10% weight loss, 25.5% with <5% weight loss, and in 20.6% patients with no weight loss or weight gain. At the individual patient level, weight change and stiffness change were not significantly correlated (Pearson r = 0.082, P = 0.165; Spearman ρ = −0.020, P = 0.732). These analyses suggest that body-weight change alone did not fully explain changes in liver stiffness observed in this cohort. In a sensitivity analysis defining follow-up weight as the minimum observed weight between 3 months after semaglutide initiation and 1 month after the follow-up elastography assessment, the association between weight change and liver stiffness change remained small (Pearson r = −0.027, P = 0.65; Spearman rho = −0.002, P = 0.98; **Table S4**).

**Fig. 6.**
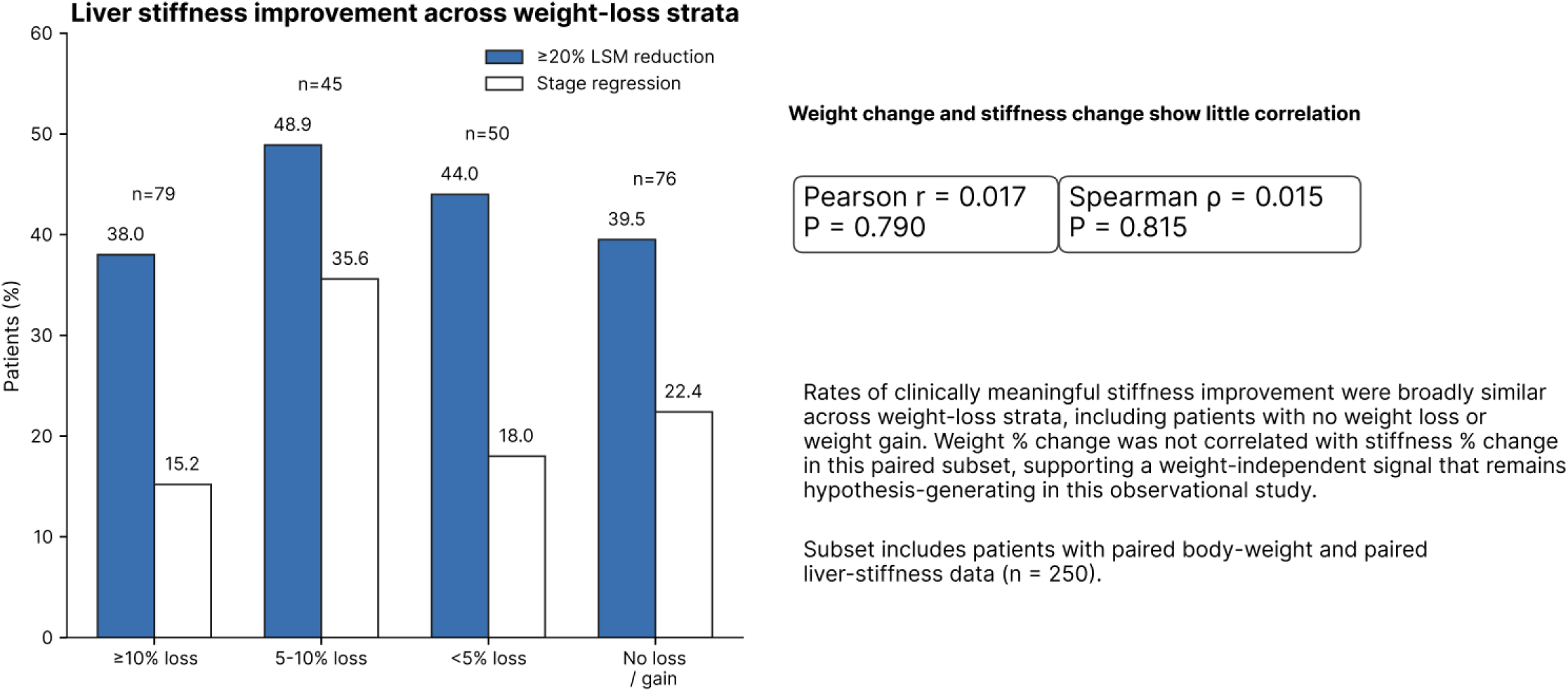
Liver stiffness improvement across weight-loss strata. Patients were stratified by percent body-weight change between selected baseline and follow-up windows. Bars show the proportion achieving >=20% stiffness reduction and stage regression. The scatter panel displays the weak correlation between percent weight change and percent stiffness change.

Semaglutide was also associated with other laboratory changes compatible with metabolic improvement, including mean reductions of HbA1c by 0.81±1.16 percentage points (p=4.5×10^−18^), ALT by 9.93±39.03 U/L (1.74×10^−11^), and AST by 4.99±33.03 U/L (p=4.71×10^−5^) (**Table S6**). However, none of these reductions were significantly correlated with change in liver stiffness, (all Spearman correlation coefficients <0.1; **Table S7**). The rates of ≥20% liver stiffness improvement were broadly similar in obese and non-obese patients (41.3% vs. 38.9%, p = 0.56), cirrhosis (42.3% vs. 38.5%, p = 0.77), dyslipidemia (42.2% vs. 37.1%, p = 0.61), or recorded MASLD-related diagnoses during the baseline period (**Table S8**). There was a trend toward a higher rate of improvement in patients without versus with type 2 diabetes (37.3% vs. 47.0%, p = 0.05), although this difference was not statistically significant. Finally, the composite FIB-4 index correlated with baseline stiffness (Spearman r = 0.66, p = 8.40×10^−30^), but FIB-4 change did not correlate with liver stiffness change (Spearman r = 0.068, p = 0.26; **Table S9**), underscoring that serial elastography captures a component of liver remodeling that is incomplete, reflected by routine serum indices alone.

Taken together, these findings suggest that while semaglutide use is associated with various cardiometabolic benefits, the observed reduction in liver stiffness does not track strongly with baseline disease diagnoses or established laboratory and anthropometric parameters.

### Single-cell GLP1R RNA expression is observed in fibrogenic and non-parenchymal liver niches

To contextualize the biological basis of the above findings, we examined GLP1R expression across 1,444,734 liver cells in public single-cell atlases (**Fig. 7**). Only 346 of 1,444,734 (0.024%) liver cells from The CZI BioHub were GLP1R-positive^13^. Hepatocytes in particular demonstrated minimal GLP1R expression, with transcript detection in only 35 of 258,760 (0.014%) individual cells. The prevalence of transcript detection was similarly low in multiple hepatocyte subsets including midzonal (3 of 19,814 cells; 0.015%), centrilobular (5 of 46,314 cells; 0.011%) and periportal hepatocytes (5 of 80,514 cells; 0.0062%). By contrast, relatively higher expression was detected in intrahepatic cholangiocytes (4 of 8,738; 0.046%; provided TEP 7.32), cholangiocytes (6 of 14,301; 0.042%; TEP 6.81), sinusoidal endothelial cells (17 of 44,583; 0.038%; TEP 6.70), and hepatic stellate cells (10 of 38,229 cells; 0.026%; TEP 4.58). GLP-1R transcript detection was also rare in immune cell populations including macrophages (16 of 125,020 cells), Kupffer cells (3 of 24,884 cells), and tissue-resident macrophages (3 of 24,895 cells). Overall, the atlas does not support abundant hepatocyte GLP1R expression and rather identifies rare GLP1R-positive cells across mesenchymal, endothelial, and biliary niches that are known to shape hepatic fibrogenesis.

**Fig. 7.**
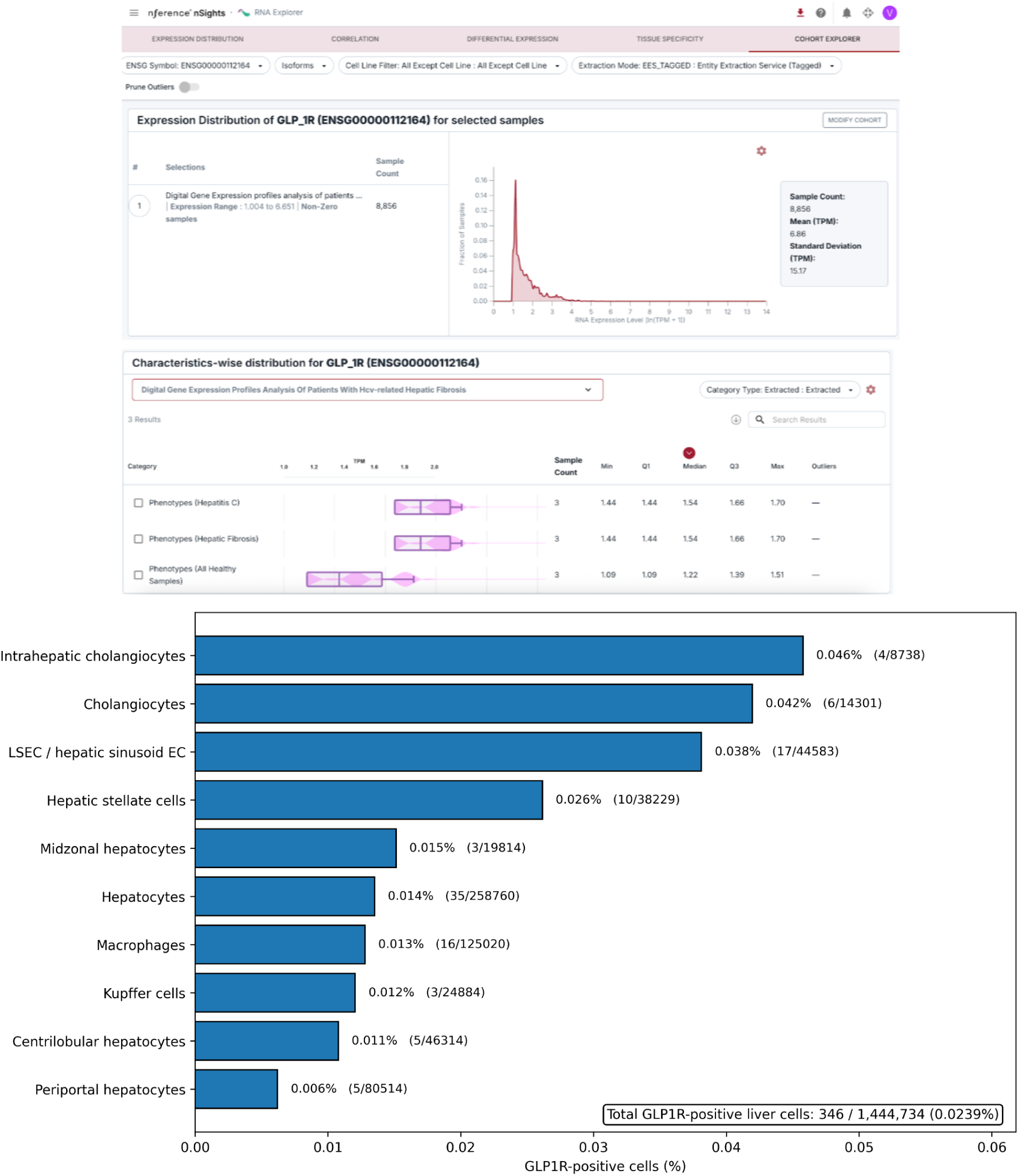
A | GLP1R bulk RNA-seq expression in hepatitis fibrosis samples and healthy liver. GLP1R is expressed higher in Hepatic fibrosis samples (n=3) relative to healthy liver samples (n=3). **B | Sparse GLP1R Single Cell RNA-seq expression of GLP1R across liver-relevant cell niches**. Horizontal bar plot summarizing the proportion of GLP1R-positive cells across selected liver-relevant cell populations from the single-cell liver atlas used in the study. Expression was sparse overall, with only 346 of 1,444,734 cells identified as GLP1R-positive (0.0239%). Relative enrichment was observed in intrahepatic cholangiocytes, cholangiocytes, liver sinusoidal endothelial cells, and hepatic stellate cells, whereas hepatocyte populations showed very low prevalence. These data support a model in which GLP1R signal in the liver is rare and localized to multicellular niches relevant to fibrogenesis, vascular remodeling, and biliary-immune crosstalk, rather than reflecting abundant hepatocyte expression.

## Discussion

In this observational routine-care analysis, higher dose semaglutide exposure is associated with a dose dependent gradient of weight loss and selective downstream hepatoprotection, most evident for steatohepatitis, alcoholic liver disease, and all cause mortality. The concordance between dose escalation, deeper weight reduction, and lower risk of steatotic and inflammatory liver phenotypes raises the possibility of adiposity loss as a principal mediator of early hepatic benefit. In contrast, the absence of signal across advanced endpoints, including cirrhosis, portal hypertensive complications, and hepatocellular carcinoma, likely reflects both limited event accrual and the longer temporal horizon required for fibrosis regression. Weight loss stratified analyses further reinforce this model, with lower risk of steatohepatitis and steatotic liver disease at higher weight loss, albeit with a non monotonic pattern suggesting heterogeneity in treatment response, adherence, or residual confounding. Collectively, these data support a framework in which higher therapeutic exposure and resultant weight loss may confer early benefit in steatotic and inflammatory liver disease, while more advanced pathology remains less modifiable within the observed timeframe.

Semaglutide treatment was associated with lower liver stiffness over time, with clinically meaningful improvement in approximately two of five patients and elastography-based stage regression in approximately one of five. The most striking reductions occurred in patients who started with the highest stiffness values, whereas patients already in the low-stiffness range changed little. That pattern is biologically and clinically plausible: patients with more advanced disease have more reversible stiffness burden, while patients near normal values have both less room to improve and greater susceptibility to floor effects.

These findings extend an evolving evidence base for semaglutide in MASH. In its 2021 phase 2 trial, semaglutide robustly increased steatohepatitis resolution but did not significantly improve fibrosis stage relative to placebo at 72 weeks^6^. A subsequent phase 2 trial in compensated MASH cirrhosis did not show a significant fibrosis benefit^7^. By contrast, the phase 3 ESSENCE interim analysis showed both MASH resolution and fibrosis improvement, findings that supported US Food and Drug Administration approval in 2025 and later AASLD practice guidance updates [8,9]. Our data add a complementary real-world view by focusing on serial elastography trajectories captured during routine care rather than trial-mandated histology. They are also directionally consistent with newer observational and secondary-analysis data linking GLP-1 receptor agonist exposure to lower cirrhosis risk, slower fibrosis progression, or improvement in liver-risk biomarkers^14–16^.

The near-flat relationship between weight change and stiffness change is one of the most interesting findings in this cohort. The simplest overinterpretation would be to call this proof of a direct liver effect; our data do not justify that claim. Residual confounding, incomplete weight capture, selective follow-up, and the imperfect link between body weight and visceral or ectopic fat loss could all obscure mediation. Nonetheless, the signal remains notable because semaglutide clearly improved weight, BMI, glycemia, and liver enzymes, yet those improvements did not discriminate stiffness responders from non-responders. In this dataset, therefore, liver stiffness improvement was not well explained by body-weight change alone. That interpretation is compatible with emerging mechanistic literature, but again only in a hypothesis-generating way. Prior research in preclinical MASH models and human proteomics showed semaglutide-linked suppression of inflammatory and fibrotic pathways^10^. At the same time, a 2024 study reported no convincing direct action of GLP-1/GIP agonism in primary human hepatocytes or hepatic stellate cells^10,11^. Our atlas analysis offers a plausible bridge between those observations: GLP1R expression was rare overall, effectively non-existent in hepatocytes, and sparse in hepatic stellate, biliary, sinusoidal endothelial, and immune niches. Together with established roles for these compartments in fibrogenic signaling networks^17–20^, this pattern supports a model in which semaglutide modifies hepatic fibrogenesis indirectly through systemic metabolic benefit.

Methodologically, this study also illustrates the value of AI-assisted extraction from longitudinal radiology text. In hepatology practice, clinically important endpoints such as elastography stiffness, CAP, technical quality metrics, and narrative interpretation often reside in free-text reports rather than harmonized analytic tables. A date-anchored agentic retrieval workflow allowed us to reconstruct these longitudinal signals at scale from a mixed structured-plus-unstructured EHR environment. That use case fits a broader movement toward AI-enabled hepatology and LLM-based radiology information extraction, especially when privacy-preserving local deployment is feasible^21–23^.

Several limitations should temper interpretation. First, this was an uncontrolled observational study, so we cannot separate semaglutide effects from concurrent lifestyle change, co-medication effects, regression to the mean, or other unmeasured confounding. Second, liver stiffness is a clinically useful but imperfect surrogate for fibrosis: it can be altered by inflammation, congestion, cholestasis, steatosis, technical factors, and our cohort included more than one elastography modality^10,11,18^. Third, most patients contributed a single pre-index and a single post-index scan, which limits trajectory modeling and makes exact timing important. Fourth, paired data for dose intensity, persistence, CAP or MRI-PDFF, and some laboratory measures were incomplete. Fifth, the cohort was heavily enriched for MASLD-related diagnoses and may not generalize to other liver disease etiologies.

Despite those caveats, the overall pattern was coherent: semaglutide-treated patients in routine practice frequently moved toward lower liver stiffness, the absolute improvements were largest in patients with more severe baseline disease, and the signal was not restricted to the largest weight losers. If confirmed in active-comparator and dose-stratified analyses with explicit manual validation of the extraction pipeline, serial elastography may provide a practical real-world endpoint for monitoring liver benefit outside clinical trials.

## Methods

### Study design and data source

We conducted a retrospective observational study using longitudinal de-identified data derived from the nSights Federated EHR Network^24–27^. The study was designed to (1) evaluate longitudinal changes in liver stiffness after initiation of semaglutide; (2) characterize concordance between changes in liver stiffness, and body weight; and (3) assess categorical transitions in fibrosis stage.

### Liver Outcomes and Landmark Analyses

Of 269,390 patients who initiated semaglutide between March 2018 and January 2024, 6,734 had baseline liver conditions and constituted the cohort examined in this observational study. Outcomes included all-cause mortality and incident liver outcomes defined using prespecified ICD-9 and ICD-10 code sets for steatotic liver disease, steatohepatitis, fibrosis, cirrhosis, portal hypertension, esophageal varices, ascites, hepatic encephalopathy, hepatorenal syndrome, spontaneous bacterial peritonitis, jaundice or cholestasis, alcoholic liver disease, viral hepatitis, autoimmune or cholestatic liver disease, drug-induced liver injury, acute liver failure, hepatocellular carcinoma, and liver transplant; complete code definitions are provided in **Table S1**. For each incident outcome analysis, patients with evidence of that same condition before follow-up were excluded. We then performed within-semaglutide 2-year landmark analyses to assess associations of attained dose intensity and achieved weight loss with subsequent liver outcomes. For dose-based analyses, the maximum semaglutide dose reached between index and the 2-year landmark was classified as low dose (0.25-1.0 mg) or high dose (>=1.7 mg), and patients were required to remain under observation through the landmark with at least one dose-bearing prescription available. For weight-loss-based analyses, baseline weight was defined as the measurement closest to index from 90 days before through 14 days after index, and maximum percent body-weight reduction was calculated from the lowest observed weight recorded from day 15 after index through the 2-year landmark. Patients were categorized into prespecified weight-loss strata of <5%, 5-10%, 10-15%, 15-20%, 20-25%, and >=25%. Post-landmark follow-up extended for 24 months and continued until the outcome of interest, death, or the last observed clinical record, whichever occurred first.

### Elastography Study Cohorts and Exposure Definition

We identified adult patients with candidate paired elastography examinations and semaglutide exposure with quantitative stiffness values available (n=326). Index date was defined as the earliest of note confirmed semaglutide initiation date or first semaglutide prescription date. All available elastography reports, including transient elastography (FibroScan) and shear-wave elastography, were identified from structured fields and unstructured clinical notes. For patients with multiple measurements within each window, the closest pre-index and latest post-index values were selected for primary analyses.

### Baseline covariates and time-aligned follow-up measurements

Baseline comorbidities, including type 2 diabetes, MASLD, obesity, cirrhosis, hypertension and dyslipidemia, were ascertained during the 10-year pre-index period. Baseline weight, BMI, HbA1c, ALT, AST and platelet count were defined as the closest available measurements within 1 year before the index date. For paired analyses, follow-up values were defined as post-index measurements obtained at least 6 months after index and nearest to the post-treatment elastography date within ±90 days. FIB-4 was calculated as age (years) × AST (U/L) / [platelet count × √ALT (U/L)]^28^. For longitudinal analyses, age at reference was held constant across baseline and follow-up FIB-4 calculations.

### Agentic Extraction of Elastography Measurements

Elastography measurements were extracted from the EHR using a large language model (LLM)-based agentic extraction workflow powered by gpt-oss-120b^29^ (temperature = 1.0, top-p = 1.0, max tokens = 64k). The agent autonomously planned and executed multi-step retrieval across both structured tables and unstructured clinical notes indexed with dense vector embeddings generated by mixedbread-ai/mxbai-embed-large-v1^30^ (see **Supplementary Methods** for system architecture and data infrastructure details).

Data extraction followed a two-phase, date-anchored retrieval strategy^31^. In Phase 1, the agent queried the structured database to identify all elastography procedures recorded for the patient, establishing a definitive set of study dates that served as temporal anchors for all downstream retrieval. In Phase 2, for each confirmed procedure date, the agent issued targeted queries to both data stores. For unstructured retrieval, semantic similarity search was performed^32^: a natural language query, enriched with clinically relevant synonyms and phrase variations to maximize recall, was encoded into a dense embedding vector and compared against the pre-indexed note chunk embeddings using cosine similarity, scoped to a date window ranging from 1 month before to 3 months after each procedure date. These chunks provided detailed measurements (stiffness in kPa, IQR, CAP, fat fraction, fibrosis stage,, acquisition quality metrics) and interpretive text not available in discrete structured fields. For drug status retrieval, the agent queried the medication table for semaglutide prescription records and additionally searched clinical notes around each scan date for references to dose changes or initiation events, reconciling structured and free-text evidence to determine the active drug and dose at the time of each study. After retrieving evidence from both sources, the agent triangulated findings: corroborating, reconciling, or flagging discrepancies between structured and unstructured data, before generating a final structured output.

The extraction prompt instructed the model to return one row per elastography study with the following fields: study date, imaging modality, device, anatomical site, stiffness (kPa), interquartile range (IQR), IQR/Median ratio, controlled attenuation parameter (CAP, dB/m), hepatic fat fraction (%), fibrosis stage (F0–F4 where documented), acquisition quality metrics, key interpretive text, active semaglutide dose at study date, drug start date, and elapsed time since drug initiation. Values not present in the source record were recorded as NA; no imputation or inference was performed. When a single session included both liver and spleen acquisitions, these were recorded as separate rows.

### LLM-Based Extraction System Architecture

Elastography and semaglutide data were extracted using a locally deployed LLM-based clinical data extraction platform built on a modular multi-tool agentic framework. The system was hosted on a Kubernetes cluster and served by a managed inference server running gpt-oss-120b with intermediate reasoning capability. The system exposes a single orchestrating agent that autonomously plans and executes multi-step extraction workflows using a set of specialized tools, with a maximum of 15 sequential reasoning turns per patient.

Prior to the full extraction, the retrieval protocol and extraction prompt were developed iteratively on a small set of representative patients using an interactive user interface. During this phase, the agent’s step-by-step reasoning and tool call sequences were observed to understand how it navigated to the relevant data, and the prompt was refined accordingly to guide the agent toward the most direct and reliable retrieval path. Once the protocol was finalized, extraction runs were executed programmatically from an analytics workspace environment by invoking the system’s REST API endpoints, enabling structured batch-style processing across the patient cohort.

### Data Infrastructure

All patient data used in this study were deidentified prior to analysis. The EHR data resided in two complementary stores. First, a structured store consisting of a columnar database organized according to a standardized common data model (CDM) schema. Clinical data spanning multiple domains were represented as discrete, timestamped events, with raw source values harmonized and vocabulary-mapped into standardized concepts, enabling consistent querying across heterogeneous source encodings. Second, an unstructured store consisting of a longitudinal corpus of full-text clinical documents spanning multiple note types (including but not limited to Clinical Notes, Radiology Notes, Pathology Notes, Lab Test Notes, and Surgical Case Notes). Documents were pre-processed into text chunks and indexed using dense vector embeddings.

### Context Management

To maintain coherent multi-turn reasoning within the model’s context window, the system applied a cascade of context compression rules when the occupied context exceeded a defined tolerance threshold: (1) removal of intermediate reasoning traces from prior turns, (2) suppression of raw tool response payloads, and (3) abstractive summarization of earlier conversation history. These rules were applied in sequence until the context size fell within tolerance, preserving the most recent turns verbatim at each stage.

### Outcomes

The primary outcome was the change in liver stiffness from pre-treatment to post-treatment elastography, where a negative delta indicated a decrease in liver stiffness. For patients with multiple studies during the study window, the closest study before index and most recent study after index during the study window were considered. Secondary outcomes included categorical transitions in fibrosis stage^33^. Stages were defined based on stiffness values as F0-F1 (<7.0 kPa), F2 (7.0-9.5 kPa), F3 (9.5-12.5 kPa), and F4/cirrhosis-range (>=12.5 kPa)^33^. Additional analyses evaluated concordance between changes in liver stiffness and body weight (Supplementary Table S3a).

### Analysis of GLP1R Expression Using Single-Cell

Single-cell expression of *GLP1R* was obtained from the CELLxGENE data portal^13^ (https://cellxgene.cziscience.com/), leveraging 1556 publicly available human single-cell and single-nucleus RNA sequencing datasets corresponding to 86 million cells from 11,633 human donors. Using the CELLxGENE Census, GLP1R expression was obtained across available cell types. Gene expression values were extracted as normalized counts (counts per 10,000 [CP10K], log-transformed where applicable) and analyzed at the cell-type level using curated annotations provided within each dataset. For each cell type, *GLP1R* expression was summarized as both mean normalized expression and the proportion of expressing cells (non-zero counts). Where multiple datasets contributed to the same tissue, results were aggregated to ensure robustness across cohorts, and analyses were restricted to cell types with adequate representation to minimize sparsity-driven artifacts.

Single-cell GLP1R atlas data were analyzed as follows. For each tissue-cell annotation, Expression (CP10K), number of GLP1R-positive cells, and percentage of expressing cells were analyzed. The primary metric was *GLP1R Engagement Potential (GEP)*, defined as Expression (CP10K) × (% expressing cells / 100). Conceptually, GEP is a prevalence-weighted expression metric: it becomes high when a cell class has both appreciable GLP1R transcript abundance and a meaningful proportion of cells that are GLP1R-positive. It is therefore useful for ranking which cellular compartments are most likely to be engaged under a broad systemic exposure model, because it balances intensity and prevalence rather than privileging either one alone. A compartment with very high expression in only a tiny number of cells may not rank as highly by GEP as a compartment with slightly lower expression but a much broader GLP1R-positive fraction.

### Statistical Analysis

Continuous variables are presented as means with standard deviations for approximately symmetric distributions and medians with interquartile ranges for skewed distributions. Categorical variables are presented as n (%). Time-to-event outcomes were evaluated using 2-year post-landmark cumulative event curves and post-landmark event proportions stratified by attained semaglutide dose and achieved weight loss. For dose-based comparisons, relative event rates were summarized as high-versus-low dose rate ratios, and differences between dose groups and across weight-loss strata were assessed using log-rank tests, with two-sided P values reported. Because liver stiffness values and paired changes were right-skewed, the primary analysis of within-patient liver stiffness change used the Wilcoxon signed-rank test; paired t-tests were used as supportive sensitivity analyses for approximately normally distributed paired differences. Correlations between change in liver stiffness and changes in weight, BMI, HbA1c, ALT, AST, and FIB-4 were assessed at the individual-patient level using Spearman rank correlation, with two-sided P values derived from the corresponding correlation test. Comparisons across baseline fibrosis severity strata used the Kruskal-Wallis test. All p values were two-sided. Statistical analyses were performed in Python 3.10.2 using pandas 2.2.2, SciPy 1.31.0, and statsmodels 0.14.2.

## Data Availability

This study involves the analysis of de-identified Electronic Health Record (EHR) data via the nference nSights Federated Clinical Analytics Platform (nSights). Data shown and reported in this manuscript were extracted from this environment using an established protocol for data extraction, aimed at preserving patient privacy. The data has been de-identified pursuant to an expert determination in accordance with the HIPAA Privacy Rule. Any data beyond what is reported in the manuscript, including but not limited to the raw EHR data, cannot be shared or released due to the parameters of the expert determination to maintain the data de-identification. The corresponding author should be contacted for additional details regarding nSights.

## De-identification and HIPAA compliance certification

Prior to analysis, all EHR data were de-identified under an expert determination consistent with the Health Insurance Portability and Accountability Act (HIPAA) Privacy Rule (45 CFR §164.514(b)(1)). The de-identification methodology employed a multi-layered transformation approach to both structured and unstructured data fields^34,35^. In structured data, direct identifiers including patient names and precise geographic locations were excluded entirely, while indirect identifiers underwent specific transformations: patient identifiers, medical record numbers, and accession numbers were replaced with one-way cryptographic hashes using confidential salts to preserve linkage across patient encounters; all dates were shifted backward by patient-specific random offsets (1–31 days) to preserve temporal relationships while obscuring exact event timing; the ZIP codes were truncated to two-digit state-level resolution; and continuous variables including age, height, weight, and body mass index were thresholded to prevent identification of extreme values (for example, ages ≥89 years transformed to ‘89+’ and BMI >40 transformed to ‘40+’). In unstructured clinical text, an ensemble de-identification system that combines attention-based deep learning models with rule-based methods achieved an estimated >99% recall for personally identifiable information (PII) detection, with detected identifiers replaced by plausible fictional surrogates^34^.

## Data Harmonization

To address heterogeneity in EHR data, we harmonized clinical variables including medications, anthropometric measurements, and diagnoses to standardized concepts. For medications, we first constructed a standardized drug concept database combining the nSights knowledge graph with RXNorm (https://www.nlm.nih.gov/research/umls/rxnorm/index.html) hierarchies to capture ingredient, brand, and dose-specific information. EHR medication records were matched using a hierarchical approach prioritizing RXNorm codes when available, followed by ingredient-level matching, and finally natural language processing and pattern matching on free-text medication orders when structured codes were absent. For anthropometric measurements (height, weight, BMI), we created a unified vocabulary from SNOMED (https://www.snomed.org/, https://athena.ohdsi.org) and LOINC (https://loinc.org/) terminologies and matched EHR measurement descriptions using standardized text matching algorithms with abbreviation expansion and synonym resolution; ambiguous mappings were resolved using OpenAI GPT-4o (https://platform.openai.com/docs/models/gpt-4o) with summary statistics as context, followed by manual verification. For diagnoses, we developed a hierarchical disease concept database from the nSights knowledge graph and matched EHR diagnosis descriptions and codes by identifying the most specific common child concept in the hierarchy. This approach enabled consistent identification of clinical entities while preserving granularity where available.

## Author contributions

A.J.V., K.M., and V.S. conceived the study. A.J.V., K.M., P.K., G.V., and A.A. designed the extraction and analytic workflows. P.K., G.V., and A.A. performed data extraction, curation, and quality review. A.J.V. and K.M. performed the statistical analyses. V.S., P.K., G.V., and K.M. drafted the manuscript. All authors interpreted the data, revised the manuscript critically for important intellectual content, and approved the final version.

## Acknowledgements

We thank the nference engineering team for the development of the nSights federated AI platform, and Patrick Lenehan for critical review and feedback on the manuscript.

## Supplementary Material

**Figure S1.**
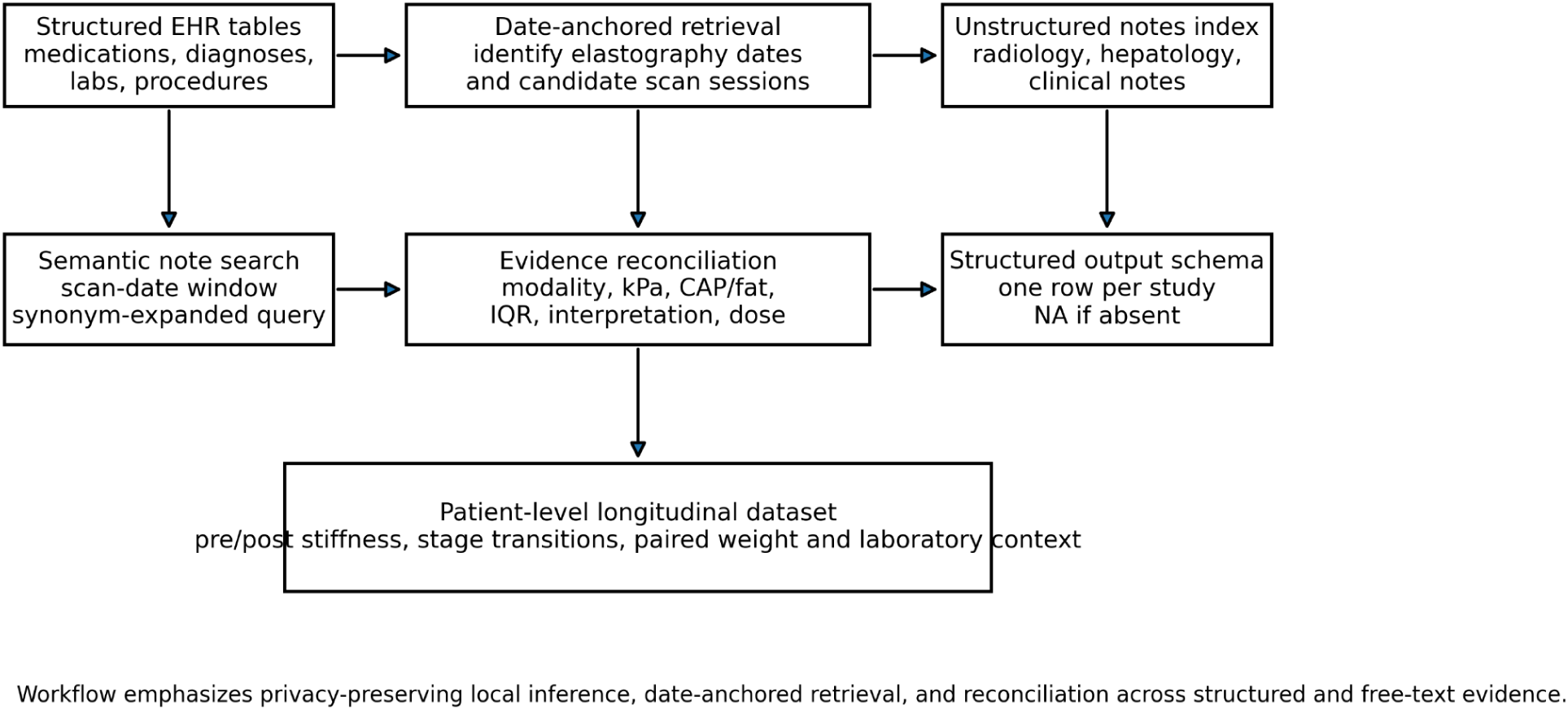
Agentic extraction workflow for longitudinal elastography and semaglutide exposure. Schematic overview of the date-anchored agentic extraction pipeline used to assemble the analytic dataset from de-identified electronic health records. Structured EHR tables were queried to identify semaglutide exposure, diagnoses, laboratory values, and procedure records, while unstructured radiology, hepatology, and clinical notes were searched using scan-date-anchored retrieval with synonym-expanded queries. Evidence from structured and free-text sources was reconciled to generate one structured output row per study session, including modality, liver stiffness, CAP/fat fraction when available, IQR and quality metrics, interpretive text, semaglutide dose, and temporal context. The resulting patient-level longitudinal dataset supported analyses of pre/post stiffness change, stage transitions, and paired weight and laboratory context.

**Fig. S2.**
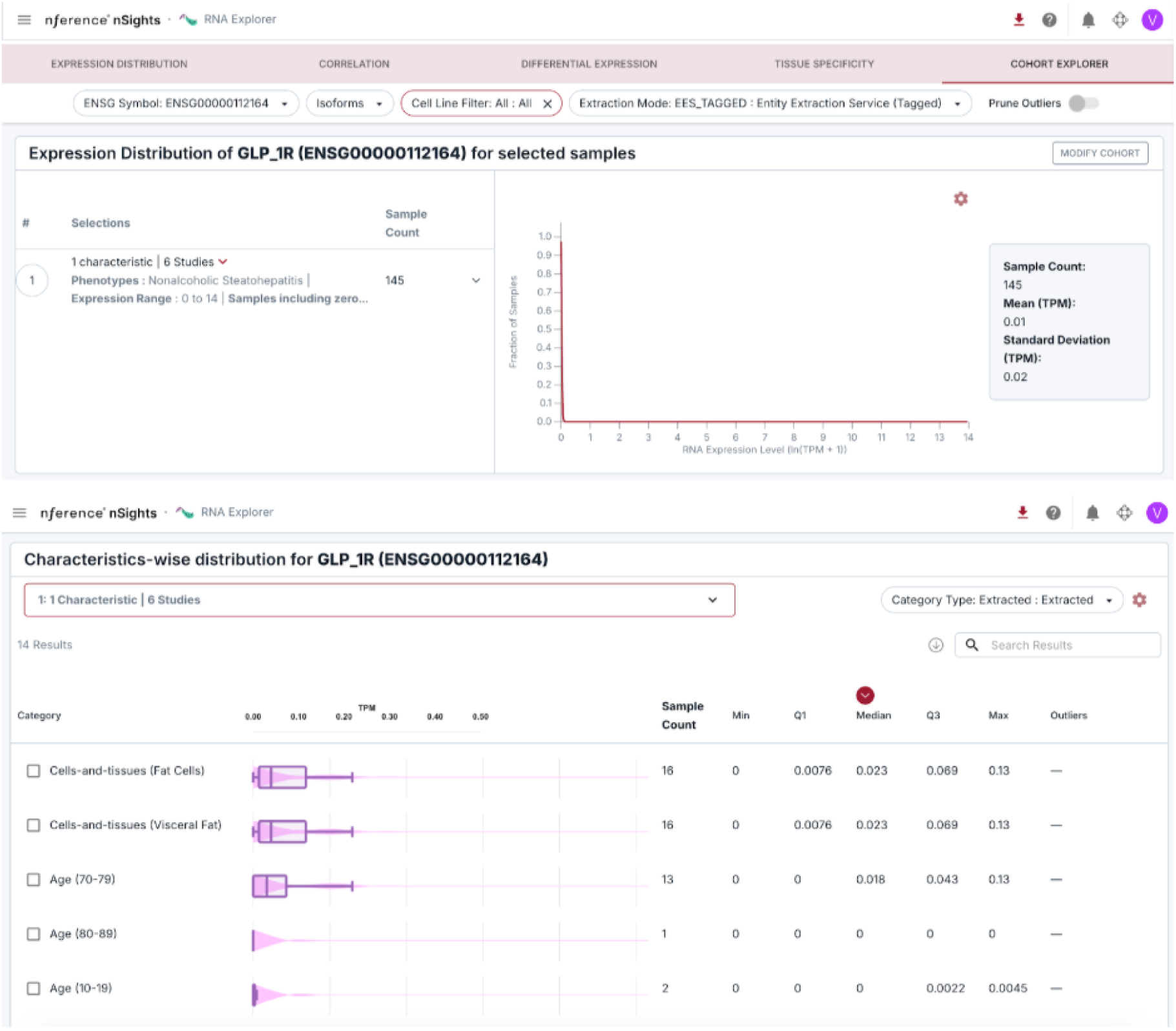
GLP1R expression in the liver from Bulk RNA-seq. **A** | GLP1R is relatively more expressed in visceral fat cells in Nonalcoholic steatohepatitis (NASH) samples (n=16) relative to other cells from NASH or healthy liver patients. https://nfer-workspaces.com/rna_lab/results?token=glp+1r&ranges=%5B0%2C0.013%2C1.004%2C6.651%5D&cell_line_data_source=all&single_cell_data_source=not_single_cell&diff_data_type=gtex_tcga&input_type=gene&collections=%5Broot-cells-and-tissues%2Croot-diseases%5D&enriched_study=high&include_cohen_d_pos=true&include_cohen_d_neg=true&single_cell_details_sort=adj_score&tissue_spec_dataset=gtex&include_cohen_d_pos_tiss_spec=true&include_cohen_d_neg_tiss_spec=true&cluster_name=all&cluster_category=all&study_exp_range=%5B0%2C14%5D&includeSamples=all&ensg_symbol=ENSG00000112164&enst_symbol=none&extraction_mode=ees_tagged&corr_data_type=whole_group&data_type=cohort_info&groupA=69d2f3672f5cc763182d7791&prune_outliers=false&isMinifyGroupA=true

## Supplementary Tables

**Table S1.** Liver phenotypes and outcomes that can be extracted pre/post semaglutide in nSights.

| Domain | Phenotype / outcome | High-yield structured codes to start with | Other allied codes / data sources to include |
| --- | --- | --- | --- |
| Steatotic liver disease | MASLD / NAFLD / fatty liver | ICD-10-CM K76.0 | SNOMED descendants; radiology/NLP terms: fatty liver, hepatic steatosis |
| Steatohepatitis | MASH / NASH | ICD-10-CM K75.81 | NLP: steatohepatitis, metabolic steatohepatitis |
| Liver fibrosis | Liver fibrosis, unspecified / other fibrosis | ICD-10-CM K74.00, K74.01, K74.02, K74.2 | Elastography kPa; pathology text; FibroScan/shear-wave report text |
| Cirrhosis | Cirrhosis, unspecified / other cirrhosis | ICD-10-CM K74.60, K74.69 | Problem list, hepatology notes |
| Portal hypertension | Portal hypertension | ICD-10-CM K76.6 | Endoscopy notes, imaging text |
| Esophageal varices | Varices without / with bleeding | ICD-10-CM I85.00, I85.01, I85.10, I85.11, I86.4 | EGD findings, procedure notes |
| Ascites | Ascites | ICD-10-CM R18.8, R18.0 | Paracentesis procedure, imaging text |
| Hepatic encephalopathy | Hepatic encephalopathy | ICD-10-CM K76.82 | Lactulose/rifaximin exposure; admission diagnoses |
| Hepatorenal syndrome | Hepatorenal syndrome | ICD-10-CM K76.7 | AKI codes, nephrology notes |
| Spontaneous bacterial peritonitis | SBP | ICD-10-CM K65.2 | Paracentesis fluid culture/cell count, inpatient notes |
| Jaundice / cholestasis | Jaundice, cholestatic disease | ICD-10-CM R17, K83.1 | Bilirubin labs, imaging text |
| Alcohol-associated liver disease | Alcoholic fatty liver / hepatitis / cirrhosis | ICD-10-CM K70.0, K70.1, K70.3x, K70.4 | Alcohol-use disorder codes, notes |
| Viral hepatitis B | Chronic HBV and sequelae | ICD-10-CM B18.1, B16.x | HBsAg, HBV DNA, antiviral meds |
| Viral hepatitis C | Chronic HCV | ICD-10-CM B18.2 | HCV RNA, antiviral meds |
| Autoimmune / cholestatic liver disease | Autoimmune hepatitis, PBC, PSC | ICD-10-CM K75.4, K74.3, K83.01 | Autoantibodies, biopsy text |
| Drug-induced liver injury | DILI / toxic liver disease | ICD-10-CM K71.x | ALT/AST spikes, causality note text |
| Acute liver injury / failure | Acute hepatic failure | ICD-10-CM K72.00, K72.01, K72.90, K72.91 | ICU/inpatient encounters |
| Hepatocellular carcinoma | HCC / primary liver cancer | ICD-10-CM C22.0 | Oncology registry, pathology |
| Liver transplant | Transplant status / history | ICD-10-CM Z94.4, Z48.23 | CPT/ICD-PCS transplant procedure history |

**Table S2.** Elastography-based fibrosis stage transition matrix.

| Baseline stage | Follow-up F0-F1 | Follow-up F2 | Follow-up F3 | Follow-up F4 |
| --- | --- | --- | --- | --- |
| F0-F1 (n=234) | 213 | 14 | 3 | 4 |
| F2 (n=39) | 27 | 6 | 4 | 2 |
| F3 (n=23) | 15 | 4 | 1 | 3 |
| F4 (n=30) | 16 | 3 | 4 | 7 |

**Supplementary Table S3.** Liver stiffness outcomes across weight-loss strata.

| Weight-loss stratum | n | >=20% stiffness improved | Stage regressed |
| --- | --- | --- | --- |
| >=10% loss | 131 | 39.7% (52) | 16.8% (22) |
| 5-10% loss | 42 | 50.0% (21) | 28.6% (12) |
| <5% loss | 47 | 38.3% (18) | 25.5% (12) |
| No loss / gain | 68 | 39.7% (27) | 20.6% (14) |

**Supplementary Table S4.**
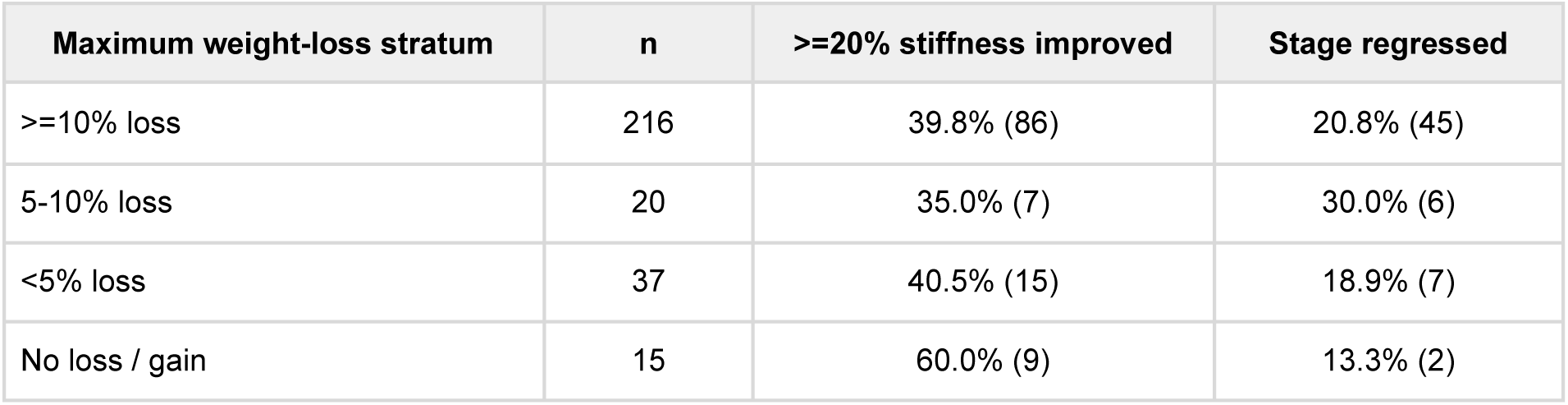
Sensitivity; Liver stiffness outcomes across maximum weight-loss strata.

**Supplementary Table S5.**
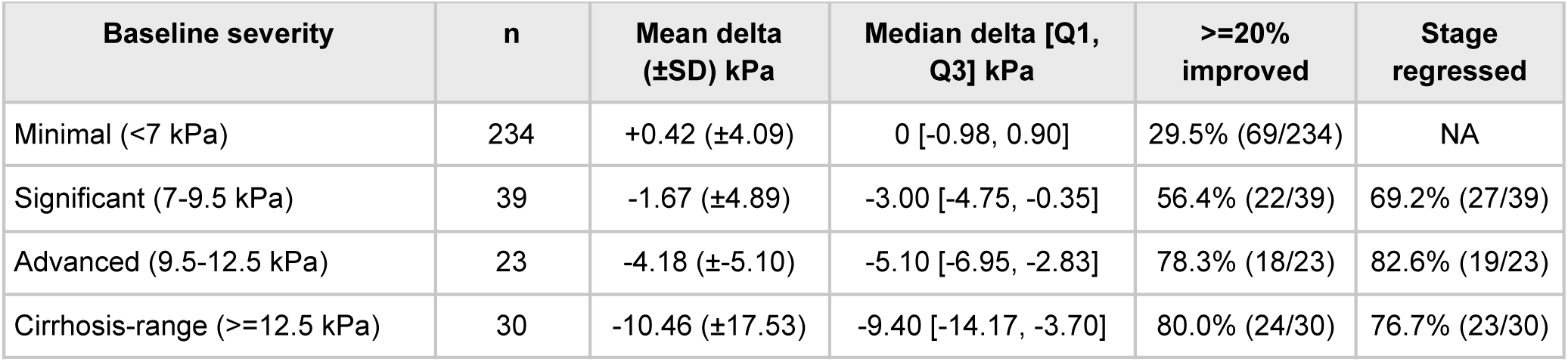
Improvement by baseline liver stiffness severity.

**Supplementary Table S6.** Paired changes in metabolic and biochemical markers.

| Marker | n paired | Pre mean | Post mean | Mean delta | t-test P | Wilcoxon P |
| --- | --- | --- | --- | --- | --- | --- |
| Weight, kg | 250 | 105.17 | 89.93 | -15.24 | 0.128 | 1.33x 10-08 |
| BMI, kg/m<sup>2</sup> | 218 | 33.37 | 31.5 | -1.87 | 8.50x 10-18 | 6.45x 10-16 |
| HbA1c, % | 158 | 6.91 | 6.1 | -0.81 | 2.25x 10-15 | 6.39x 10-17 |
| ALT, U/L | 260 | 47.52 | 37.58 | -9.93 | 5.43x 10-05 | 4.05x 10-09 |
| AST, U/L | 259 | 38.67 | 33.68 | -4.99 | 0.0157 | 1.05x 10-05 |

**Supplementary Table S7.** Correlation of metabolic changes with liver stiffness change.

| Variable | n | Pearson correlation with stiffness change | Spearman correlation with stiffness change |
| --- | --- | --- | --- |
| Weight % change | 250 | 0.017 (P=0.79) | 0.015 (P=0.815) |
| BMI % change | 218 | 0.094 (P=0.166) | 0.039 (P=0.57) |
| HbA1c delta | 158 | -0.045 (P=0.571) | 0.024 (P=0.76) |
| ALT % change | 260 | 0.011 (P=0.854) | -0.019 (P=0.761) |
| AST % change | 259 | 0.083 (P=0.186) | -0.002 (P=0.976) |

**Supplementary Table 8.** Selected subgroup analyses.

| Subgroup | n | >=20% improved | P value |
| --- | --- | --- | --- |
| Type 2 diabetes present | 209 | 37.3% (78/209) | 0.051 |
| Type 2 diabetes absent | 117 | 47.0% (55/117) |  |
| Obesity present | 254 | 41.3% (105/254) | 0.561 |
| Obesity absent | 72 | 38.9% (28/72) |  |
| Cirrhosis present | 196 | 42.3% (83/196) | 0.767 |
| Cirrhosis absent | 130 | 38.5% (50/130) |  |
| Dyslipidemia present | 237 | 42.2% (100/237) | 0.611 |
| Dyslipidemia absent | 89 | 37.1% (33/89) |  |
| Hypertension present | 253 | 40.3% (102/253) | 0.590 |
| Hypertension absent | 73 | 42.5% (31/73) |  |
| NAFLD present | 225 | 42.2% (95/225) | 0.541 |
| NAFLD absent | 101 | 37.6% (38/101) |  |

**Supplementary Table S9.** FIB-4 and liver stiffness concordance.

| **FIB-4 metric** | **Value** |
| --- | --- |
| Paired FIB-4 cohort | 231 |
| Baseline mean FIB-4 | 1.73 |
| Follow-up mean FIB-4 | 1.72 |
| Mean delta | -0.01 |
| Paired t-test P value | 0.824 |
| Wilcoxon P value | 0.893 |
| Correlation with stiffness delta (Pearson) | -0.060 (P=0.365) |
| Correlation with stiffness delta (Spearman) | +0.068 (P=0.303) |
| Baseline correlation with baseline stiffness (Pearson) | +0.567 (P=1.24 x 10<sup>-20</sup>) |
| Baseline correlation with baseline stiffness (Spearman) | +0.661 (P=8.40 x 10<sup>-30</sup>) |

## References

1. Huang, D. Q. et al. Metabolic dysfunction-associated steatotic liver disease in adults. Nat Rev Dis Primers 11, 14 (2025).

2. Younossi, Z. M., Kalligeros, M. & Henry, L. Epidemiology of metabolic dysfunction-associated steatotic liver disease. Clin Mol Hepatol 31, S32–S50 (2025).

3. Hagström, H., Shang, Y., Hegmar, H. & Nasr, P. Natural history and progression of metabolic dysfunction-associated steatotic liver disease. Lancet Gastroenterol Hepatol 9, 944–956 (2024).

4. European Association for the Study of the Liver (EASL), European Association for the Study of Diabetes (EASD) & European Association for the Study of Obesity (EASO). EASL-EASD-EASO Clinical Practice Guidelines on the management of metabolic dysfunction-associated steatotic liver disease (MASLD). J Hepatol 81, 492–542 (2024).

5. Website. https://www.fda.gov/news-events/press-announcements/fda-approves-first-treatment-patients-liver-scarring-due-fatty-liver-disease.

6. Newsome, P. N. et al. A Placebo-Controlled Trial of Subcutaneous Semaglutide in Nonalcoholic Steatohepatitis. N Engl J Med 384, 1113–1124 (2021).

7. Loomba, R. et al. Semaglutide 2·4 mg once weekly in patients with non-alcoholic steatohepatitis-related cirrhosis: a randomised, placebo-controlled phase 2 trial. Lancet Gastroenterol Hepatol 8, 511–522 (2023).

8. Sanyal, A. J. et al. Phase 3 Trial of Semaglutide in Metabolic Dysfunction-Associated Steatohepatitis. N Engl J Med 392, 2089–2099 (2025).

9. Website. https://www.fda.gov/drugs/news-events-human-drugs/fda-approves-treatment-serious-liver-disease-known-mash.

10. Bansal, M. B. et al. Semaglutide therapy for metabolic dysfunction-associated steatohepatitis: November 2025 updates to AASLD Practice Guidance. Hepatology (2025) doi:10.1097/HEP.0000000000001608.

11. Jara, M. et al. Modulation of metabolic, inflammatory and fibrotic pathways by semaglutide in metabolic dysfunction-associated steatohepatitis. Nat Med 31, 3128–3140 (2025).

12. Petta, S. et al. Monitoring Occurrence of Liver-Related Events and Survival by Transient Elastography in Patients With Nonalcoholic Fatty Liver Disease and Compensated Advanced Chronic Liver Disease. Clin Gastroenterol Hepatol 19, 806–815.e5 (2021).

13. CZI Cell Science Program et al. CZ CELLxGENE Discover: a single-cell data platform for scalable exploration, analysis and modeling of aggregated data. Nucleic Acids Res 53, D886–D900 (2025).

14. da Silva Lima, N., et al. GLP-1 and GIP agonism has no direct actions in human hepatocytes or hepatic stellate cells. Cell Mol Life Sci 81, 468 (2024).

15. Kanwal, F. et al. GLP-1 Receptor Agonists and Risk for Cirrhosis and Related Complications in Patients With Metabolic Dysfunction-Associated Steatotic Liver Disease. JAMA Intern Med 184, 1314–1323 (2024).

16. Choi, J. et al. GLP-1RA and Liver Fibrosis Progression in MASLD and Type 2 Diabetes: Target Trial Emulation Using Propensity Score Matching. Liver Int 45, e70447 (2025).

17. Meyhöfer, S. M. et al. Semaglutide on liver fibrosis and heart outcomes in patients at high risk of liver fibrosis: a prespecified analysis of the SELECT randomized trial. Nat Med (2026) doi:10.1038/s41591-026-04281-1.

18. Gawrieh, S. et al. Increases and decreases in liver stiffness measurement are independently associated with the risk of liver-related events in NAFLD. J Hepatol 81, 600–608 (2024).

19. Lin, H. et al. Vibration-Controlled Transient Elastography Scores to Predict Liver-Related Events in Steatotic Liver Disease. JAMA 331, 1287–1297 (2024).

20. Tsuchida, T. & Friedman, S. L. Mechanisms of hepatic stellate cell activation. Nat Rev Gastroenterol Hepatol 14, 397–411 (2017).

21. Qu, J., Wang, L., Li, Y. & Li, X. Liver sinusoidal endothelial cell: An important yet often overlooked player in the liver fibrosis. Clin Mol Hepatol 30, 303–325 (2024).

22. Ceci, L., Gaudio, E. & Kennedy, L. Cellular Interactions and Crosstalk Facilitating Biliary Fibrosis in Cholestasis. Cell Mol Gastroenterol Hepatol 17, 553–565 (2024).

23. Reichenpfader, D., Müller, H. & Denecke, K. A scoping review of large language model based approaches for information extraction from radiology reports. NPJ Digit Med 7, 222 (2024).

24. Venkatakrishnan, A. J. et al. Clinical nSights: A software platform to accelerate real world oncology analyses. Journal of Clinical Oncology (2024) doi:10.1200/JCO.2024.42.16_suppl.e23316.

25. Venkatakrishnan, A. J., Murugadoss, K. & Soundararajan, V. Weight Loss Dynamics and Health Burden Changes with Tirzepatide versus Semaglutide. medRxiv 2025.11.30.25341294 (2025) doi:10.64898/2025.11.30.25341294.

26. Murugadoss, K., Varma, G., Venkatakrishnan, A. J., Gibson, M. C. & Soundararajan, V. Weight trajectories after last Tirzepatide or Semaglutide prescription across a federated health network. medRxiv 2026.01.26.26344839 (2026) doi:10.64898/2026.01.26.26344839.

27. Venkatakrishnan, A. J., Murugadoss, K. & Soundararajan, V. Decoding the hallmarks of GLP-1RA weight-loss super responders. medRxiv 2025.11.15.25340314 (2025) doi:10.1101/2025.11.15.25340314.

28. Vallet-Pichard, A. et al. FIB-4: An inexpensive and accurate marker of fibrosis in HCV infection. comparison with liver biopsy and fibrotest. Hepatology 46, 32–36 (2007).

29. OpenAI et al. gpt-oss-120b & gpt-oss-20b Model Card. (2025).

30. Open Source Strikes Bread - New Fluffy Embedding Model. Mixedbread https://www.mixedbread.com/blog/mxbai-embed-large-v1.

31. Hripcsak, G. & Albers, D. J. Next-generation phenotyping of electronic health records. J Am Med Inform Assoc 20, 117–121 (2013).

32. Lewis, P., et al. Retrieval-Augmented Generation for Knowledge-Intensive NLP Tasks. (2020).

33. Castéra, L. et al. Prospective comparison of transient elastography, Fibrotest, APRI, and liver biopsy for the assessment of fibrosis in chronic hepatitis C. Gastroenterology 128, 343–350 (2005).

34. Murugadoss, K. et al. Building a best-in-class automated de-identification tool for electronic health records through ensemble learning. Patterns (N Y) 2, 100255 (2021).

35. Murugadoss, K. et al. Scaling text de-identification using locally augmented ensembles. medRxiv 2024.06.20.24308896 (2024) doi:10.1101/2024.06.20.24308896.

